# Neuroimaging correlates of symptom burden and functional recovery following mild traumatic brain injury: A systematic review

**DOI:** 10.1101/2025.04.28.25326598

**Authors:** Joshua P. McGeown, Mangor Pedersen, Remika Mito, Alice Theadom, Jerome J. Maller, Paul Condron, Samantha J. Holdsworth

## Abstract

**Background:** Mild traumatic brain injury (mTBI) represents 95% of all traumatic brain injuries. Despite being classified as “mild,” mTBI can lead to significant, long-term symptoms that impact quality of life. Diagnostic and management strategies rely heavily on subjective symptom reporting due to a lack of validated biomarkers. Identifying neuroimaging biomarkers to study the pathophysiological features contributing to symptom burden and poor recovery is critical for improving mTBI management.

**Objective:** To evaluate the relationships between acute Magnetic Resonance Imaging (MRI) findings and mTBI symptom burden and functional recovery, and to understand how these relationships evolve over time.

**Methods:** The review followed the Preferred Reporting Items for Systematic Reviews and Meta-Analyses (PRISMA) guidelines. Systematic searches were conducted in MEDLINE, SCOPUS, and Cochrane Library databases to identify studies on mTBI with acute MRI data, symptom burden or functional recovery assessments, and at least one follow-up clinical timepoint, covering publications from inception to February 5, 2024.

**Results:** Fifty-two of 6,759 articles were included. The review identified heterogeneous evidence across MRI modalities. Structural MRI findings showed limited correlation with clinical outcomes, while changes in white matter and functional connectivity were more strongly associated with symptom burden and recovery. Disruptions in multi-functional hubs such as the thalamus, superior longitudinal fasciculus, and cingulate cortex were linked to increased symptom burden and poorer recovery outcomes.

**Conclusions:** Acutely acquired MRI and clinical data provide crucial insights into the complexities of mTBI symptomology and recovery. This underscores the need to incorporate symptom burden and recovery measures in mTBI neuroimaging studies.

**Key findings:**

- Divergent neuroimaging findings in mTBI patients with incomplete vs. full recovery
- Thalamus, SLF, and cingulate disruptions linked to mTBI symptom burden and recovery
- Opportunity to guide neuroimaging analysis by clinically driven phenotypes
- Opportunity to embrace multimodal analytical frameworks to advance understanding
- Significant methodological heterogeneity across studies

## 1. Introduction

Mild traumatic brain injury (mTBI) represents 95% of all traumatic brain injuries (Feigin et al. 2013, James et al. 2019). The term “concussion” is often used interchangeably with mTBI, and other times as a distinct form of brain injury. There is now consensus that concussion is a form of mTBI (Patricios et al. 2023, Silverberg et al. 2023). Further, in 2018, a statement from the Centers for Disease Control and Prevention recommended using mTBI as the standard terminology to avoid misinterpretation (Lumba-Brown et al. 2018). In line with this recommendation, the term mTBI will be used in the remainder of the paper. While classified as “mild” because they are generally not life-threatening and do not typically result in coma or permanent disability (Sussman et al. 2018), up to 50% of individuals with such injuries may experience debilitating symptoms and a reduced quality of life 6-12 months after the injury (Feigin et al. 2013, Nelson et al. 2019, Voormolen et al. 2020). Previous research has identified several factors that predict delayed recovery from mTBI, including a pre-existing history of mental health issues or migraines, a higher acute symptom burden, female sex, delayed presentation to a clinician, and/or repeated mTBI (Zemek et al. 2016, Iverson et al. 2017, Emery et al. 2021). However, due to a lack of clinically validated objective biomarkers, the pathophysiological mechanisms contributing to the heterogeneity in mTBI symptom severity and functional recovery outcomes are poorly understood (Silverberg et al. 2020). This means that diagnosing, managing, and determining recovery from mTBI relies entirely on clinical assessment, with subjective symptom reporting playing a crucial role in clinical decision-making (Silverberg et al. 2020). One of the difficulties with this current approach is that subjective reporting can be influenced by contextual factors such as downplaying the injury to return to sport (Meier et al. 2015b, Clark and Stanfill 2019). Additionally, emerging evidence suggests that physiological recovery takes longer than the person noticeably experiences symptoms, possibly resulting in people returning to risk activities before the brain fully recovers (Kamins et al. 2017). This highlights the need for objective markers of injury and recovery following mTBI.

Kenzie et al. developed a causal-loop model of mTBI that integrates biomechanical studies, basic science, and clinical evidence. This model illustrates how mTBI induces a pathophysiological cellular environment, which impairs neurological networks and drives symptom complaints and quality of life deficits (Kenzie et al. 2018). The framework provides targets for developing potentially translatable fluid and neuroimaging biomarkers to enhance our understanding of mTBI and improve clinical outcomes for patients. Modern Magnetic Resonance Imaging (MRI) sequences offer promising methods to non-invasively investigate the pathophysiological consequences of mTBI on brain structure and function in patient populations. T1-weighted (T1w) images assess changes in brain structure and volume, while T2-weighted (T2w) and FLuid Attenuated Inversion Recovery (FLAIR) images reveal tissue abnormalities. Susceptibility Weighted Imaging (SWI) and Quantitative Susceptibility Mapping (QSM) allow for the assessment of microbleeds and iron deposition. Functional connectivity can be measured by correlating blood oxygenation level dependent (BOLD) fluctuations using resting-state functional MRI (rs-fMRI). Diffusion-weighted imaging, including Diffusion Tensor Imaging (DTI), facilitates the investigation of altered white matter structure, with more advanced forms such as Diffusion Kurtosis Imaging (DKI) allows one to investigate more complex microstructural changes. Finally, Arterial Spin Labelling (ASL) can map perfusion parameters such as cerebral blood flow (CBF) and cerebral blood volume.

Previous efforts to study mTBI using MRI have largely focused on comparing participants with mTBI against healthy controls. Systematic reviews have highlighted conflicting results across DTI (Asken et al. 2018, Hellewell et al. 2021, Jain et al. 2021, Lindsey et al. 2021), rs-fMRI (Morelli et al. 2021), and ASL (Wang et al. 2023) studies, attributing these inconsistencies to variations in study design, time since mTBI, imaging parameters, image quality, and analysis methods. Many studies, however, treat mTBI samples as a homogeneous group despite documented heterogeneity in symptomology and functional recovery outcomes. Furthermore, studies commonly group acute (0 to 14 days post-injury), subacute (14 days to 8 weeks post-injury), and chronic (12+ weeks post-injury) mTBI in the same sample for analysis. Finally, reviews to date have focused on a single MRI modality to investigate the effects of mTBI. The multifaceted nature of mTBI suggests that valuable insights may be gained from curating evidence to explore the overlapping nature of MRI findings across multiple modalities.

Given the complexity of mTBI, MRI findings may vary between patients with different levels of symptom severity or between those whose symptoms spontaneously resolve versus those who develop persistent symptoms. To better understand how MRI can provide insights into mTBI symptoms and functional recovery, collating, integrating, and synthesising the available evidence across various MRI modalities is essential. Therefore, this systematic review addresses these gaps by assessing how acute MRI findings correlate with mTBI symptom burden and functional recovery, and how these relationships evolve over time.

## 2. Methods

This review was conducted per the Preferred Reporting Items for Systematic Reviews and Meta-Analyses (PRISMA) guidelines. A priori criteria of this review were registered with the International Prospective Register of Systematic Reviews (PROSPERO; CRD42022338069), and the review methodology conformed to Population, Intervention, Comparator, and Outcome (P.I.C.O.) guidelines. Note, studies reporting MRI findings and neurocognitive outcomes without measurement of symptom burden and/or functional recovery outcomes were beyond the scope of this review.

For the purposes of this review – and based on common characteristics across various definitions (Carroll et al. 2004, Carney et al. 2014, Patricios et al. 2023, Silverberg et al. 2023) – mTBI is used to describe brain injuries with the following features:

- A plausible mechanism of injury involving the transfer of direct or indirect external forces to the brain;
- Clinical signs such as Glasgow Coma Scale (GCS; (Jain and Iverson 2020)) score of 13-15, loss of consciousness ≤30 minutes, post-traumatic amnesia ≤24 hours, gross motor instability, and/or seizure;
- Acute alteration of mental status (i.e. confusion, disorientation, dazed);
- Acute physical (i.e. headache, nausea, balance problems, photophobia), cognitive (memory problems, feeling slowed down, difficulty concentrating), or emotional (i.e. uncharacteristic emotional lability or irritability) symptoms that may present immediately or evolve within 72 hours post-injury;
- Absence of gross intracranial damage on CT or MRI (if acquired) requiring neurosurgical intervention consistent with moderate or severe traumatic brain injury;
- Signs and symptoms are not better explained by a confounding factor (i.e. psychological trauma, alcohol or substance consumption, circulatory disruption).

### 2.1 Search strategy and study selection criteria

Table 1 provides the search terms used to identify studies investigating the relationships between functional recovery outcomes following mTBI and acute/follow-up MRI findings. Using the AND operator, we filtered titles, abstracts, and keywords by combining mTBI-related, imaging-related, and modality-related search terms. Additional filtering was applied, when possible, to identify full text original research journal articles written in English. The systematic search returned 8,039 articles (Figure 1) available online and published from inception through to February 5, 2024, in MEDLINE [EBSCO] (n = 4,195), SCOPUS (n = 3,731), and Cochrane Library [Wiley] (n = 113). After removal of duplicates, the titles and abstracts of 6,759 studies were blindly screened for inclusion by JPM and RM using Rayyan Systematic Review Software. Blinding was temporarily removed after title and abstract screening to resolve any disagreements about studies requiring full-text screening for eligibility. Blinding was reactivated for full-text screening, and again, disagreements were resolved via discussion.

**Figure 1.**
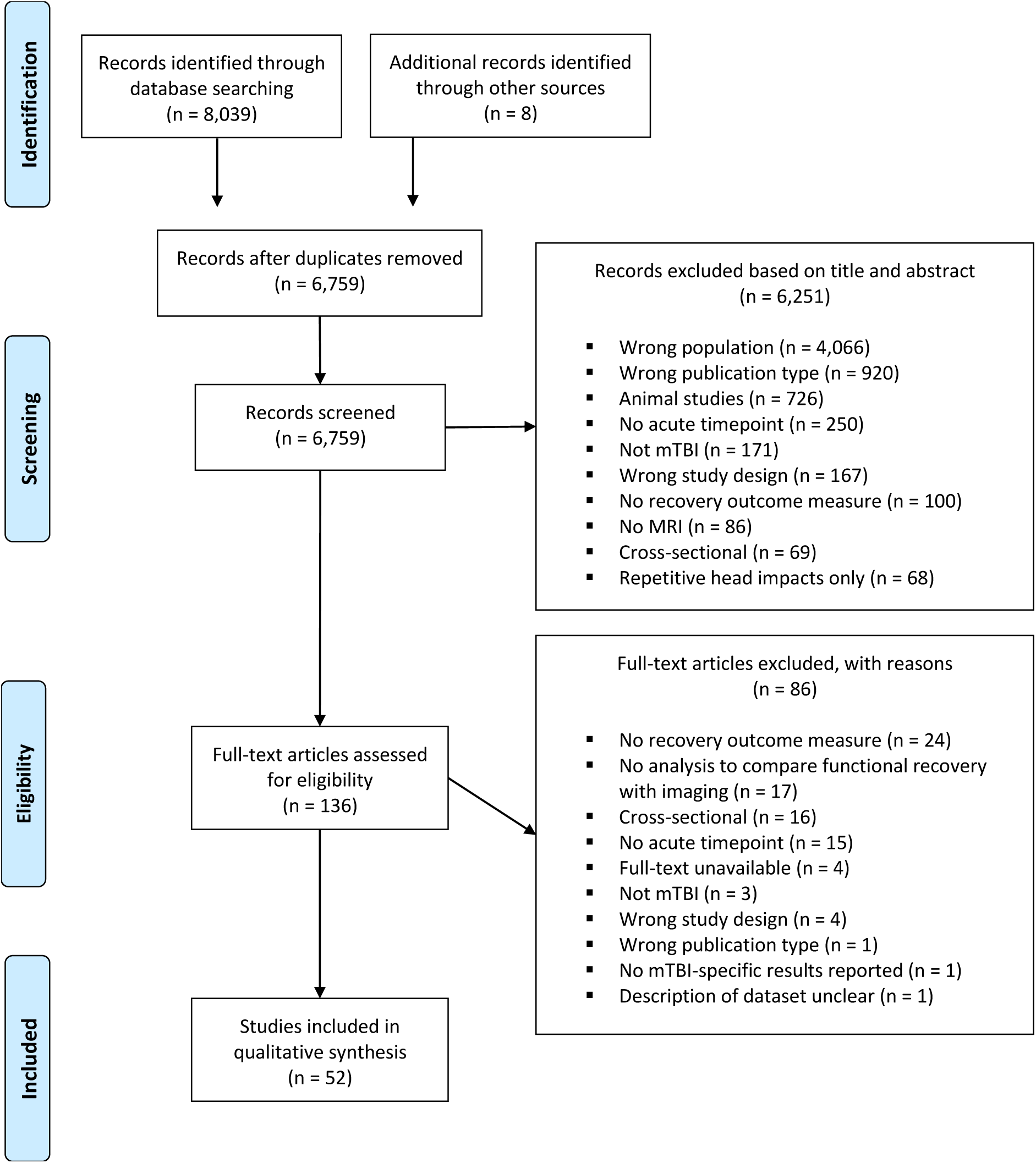
PRISMA flowchart.

**Table 1.**
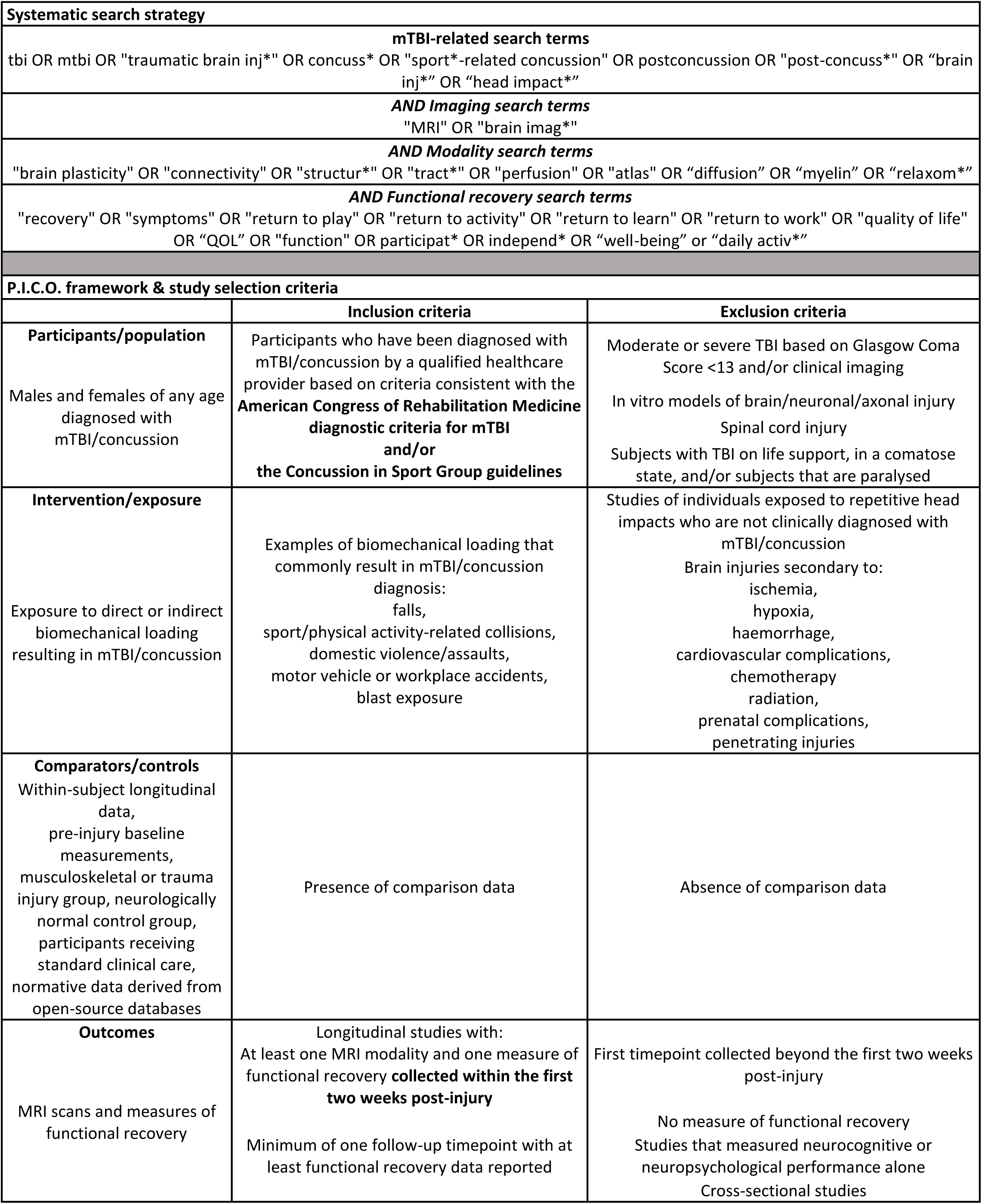
Search strategy and inclusion/exclusion criteria.

### 2.2 Data extraction

For each study, the mTBI definition, study design, sample size and characteristics, MRI sequence(s), scanner type, imaging timepoints, measures of mTBI symptoms and/or functional recovery, and results relating imaging findings to symptom/recovery findings were extracted and reported. Only significant results after multiple comparisons correction were extracted, unless otherwise indicated.

## 3. Results

Screening of titles and abstracts resulted in 130 articles assessed for eligibility. During full-text screening, a search of reference lists identified eight further studies for consideration. In total, 52 studies were included in the qualitative synthesis for this review.

### 3.1 Study characteristics

A description of the mTBI definition used, mTBI and control cohort details, MRI sequences and scanners, and whether multi-site data was used is provided for each study in Table 2.

**Table 2.**
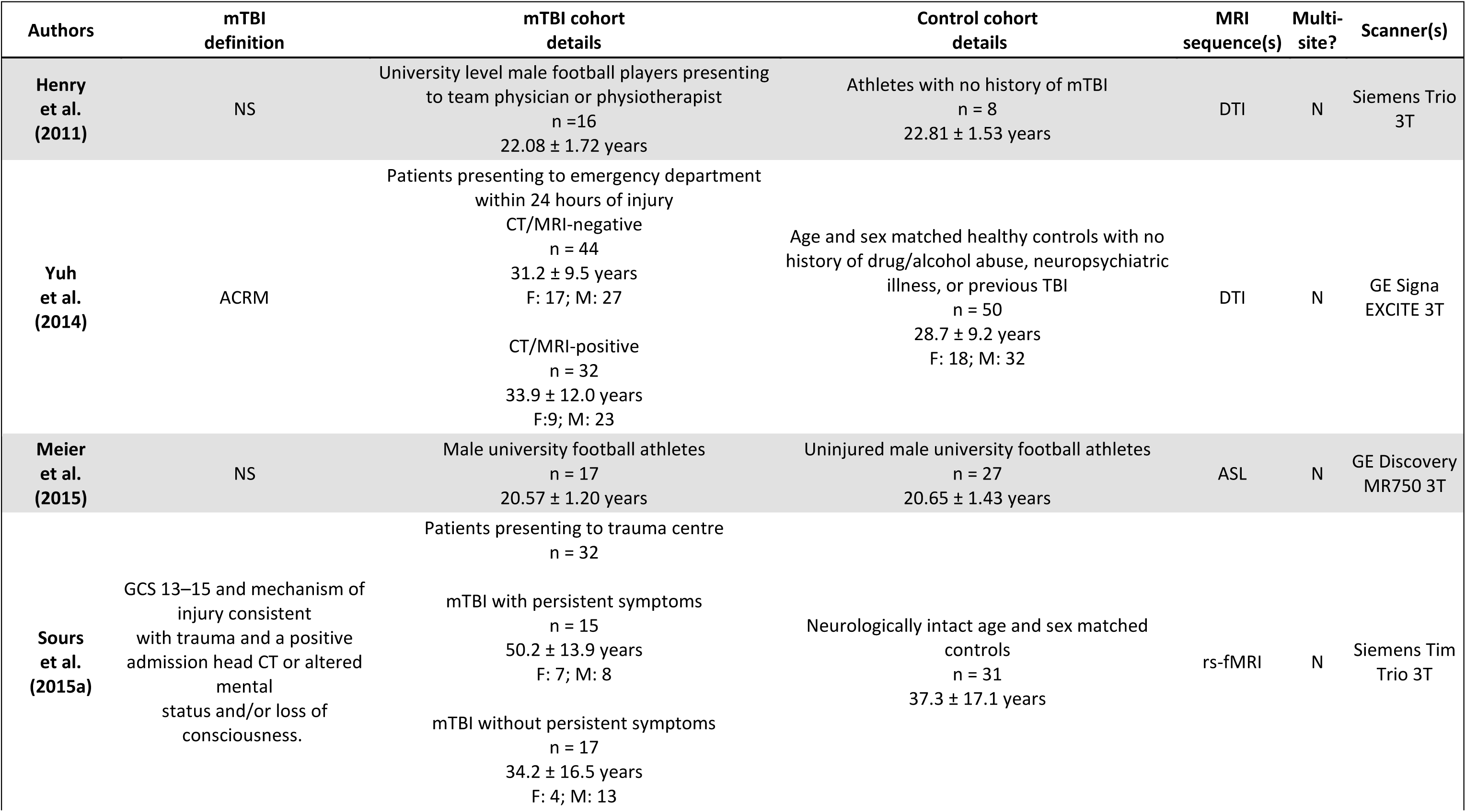
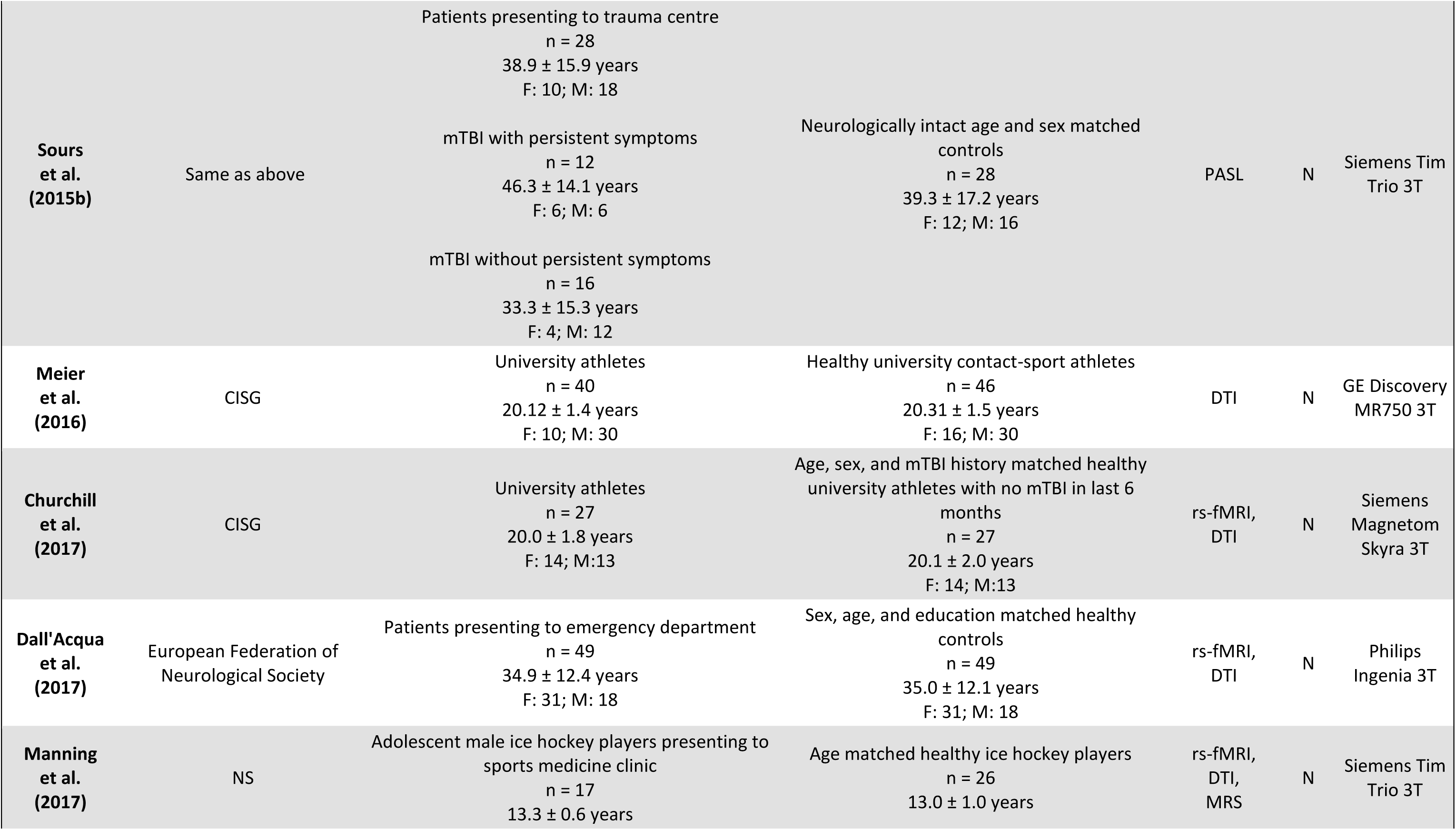
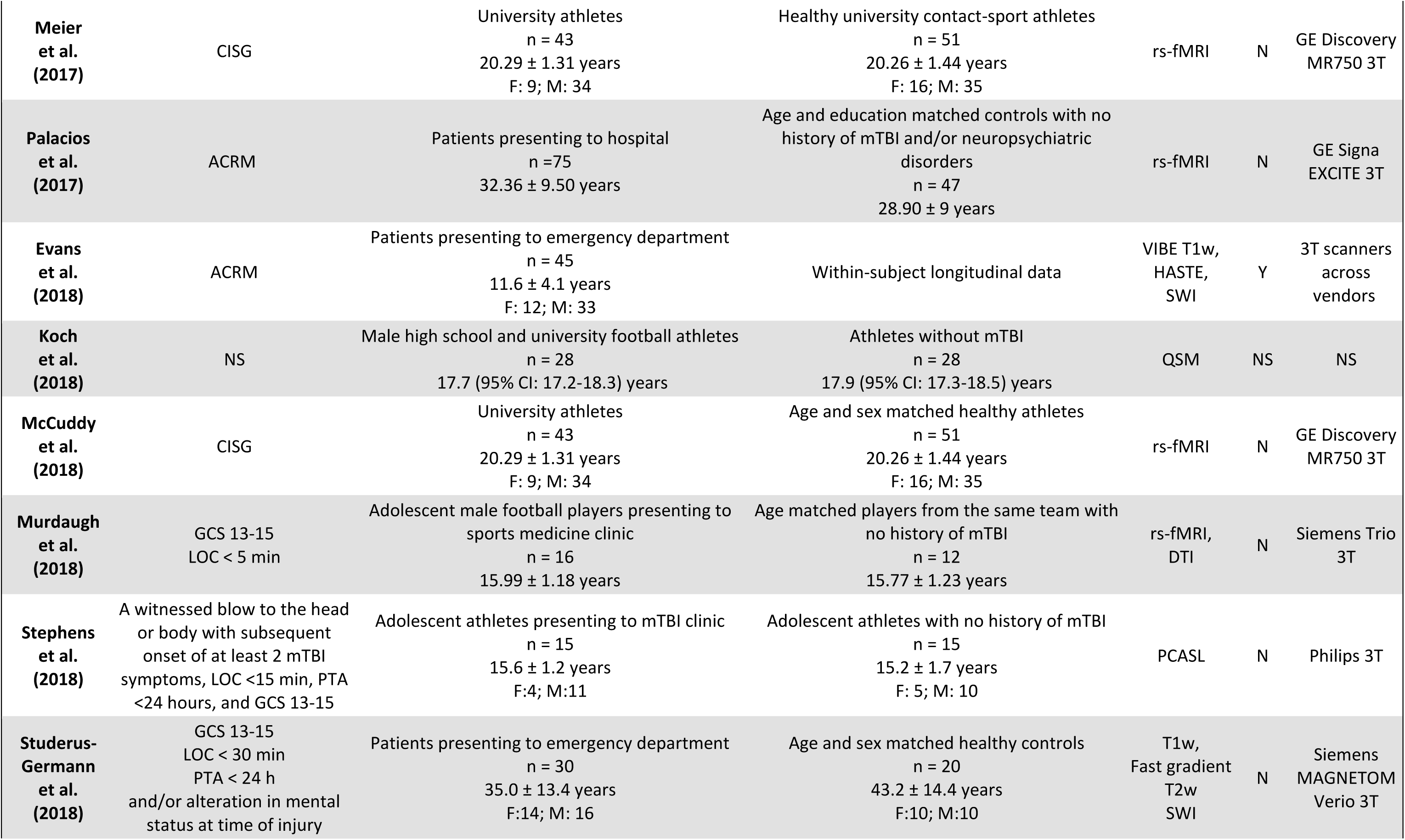
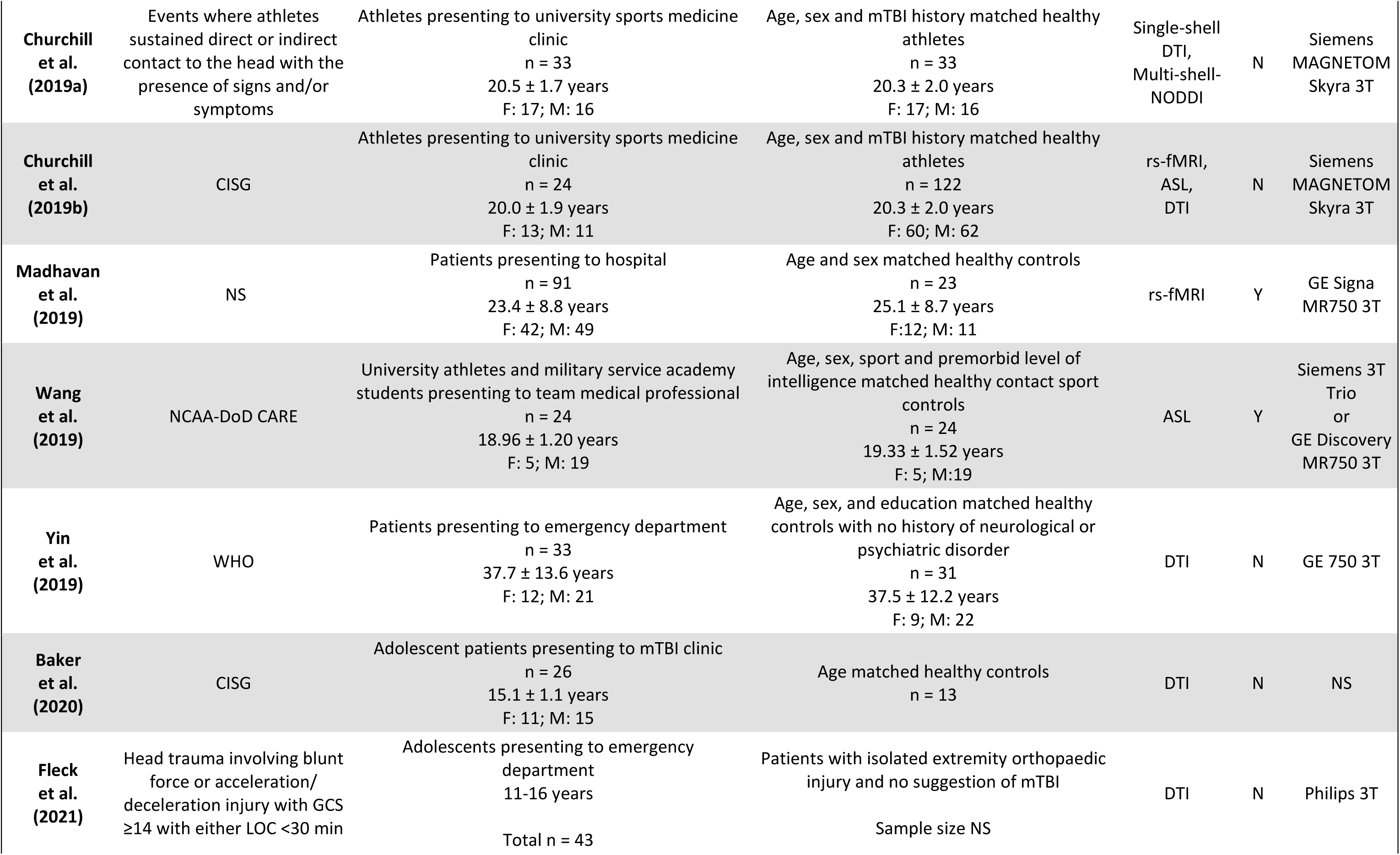
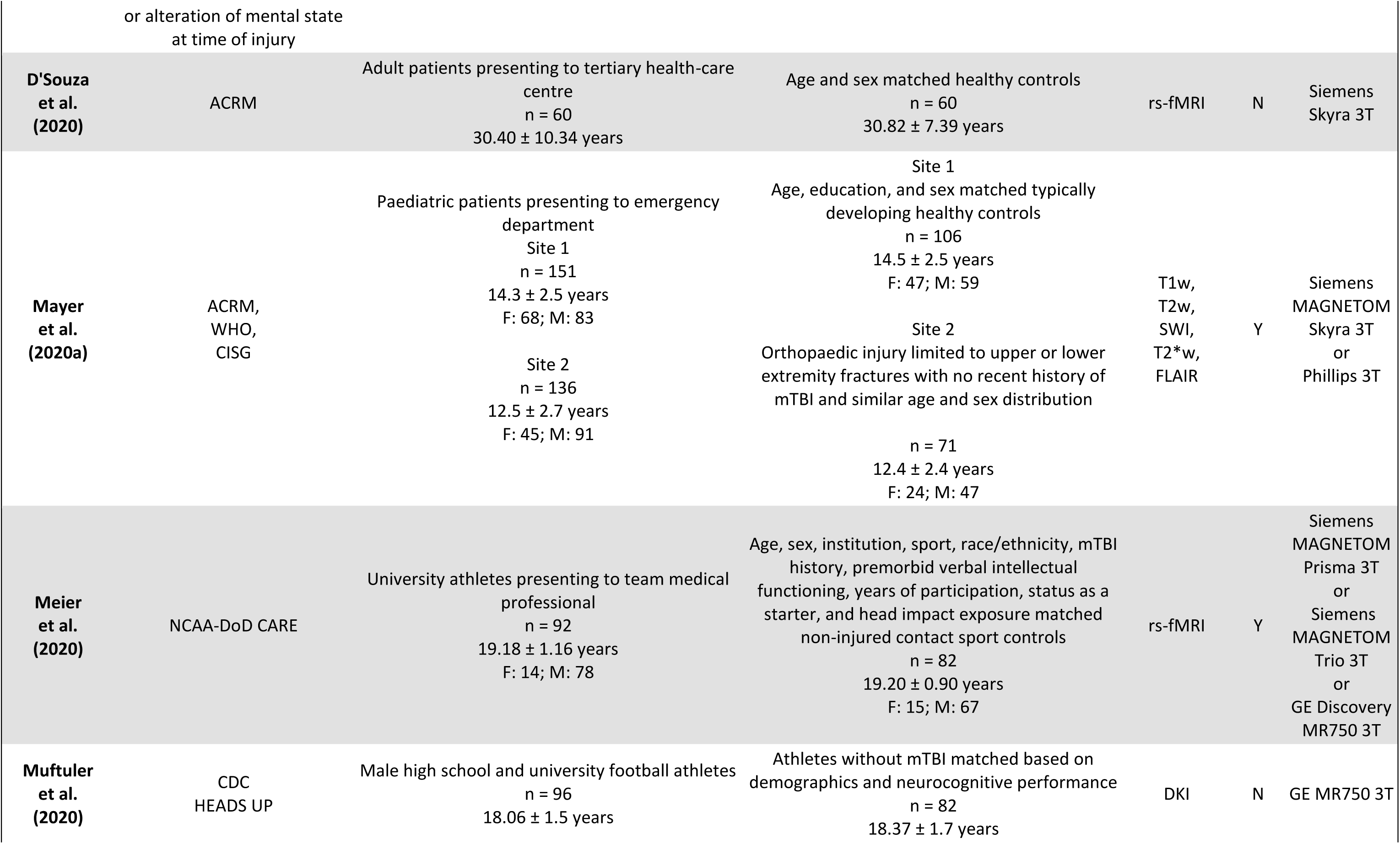
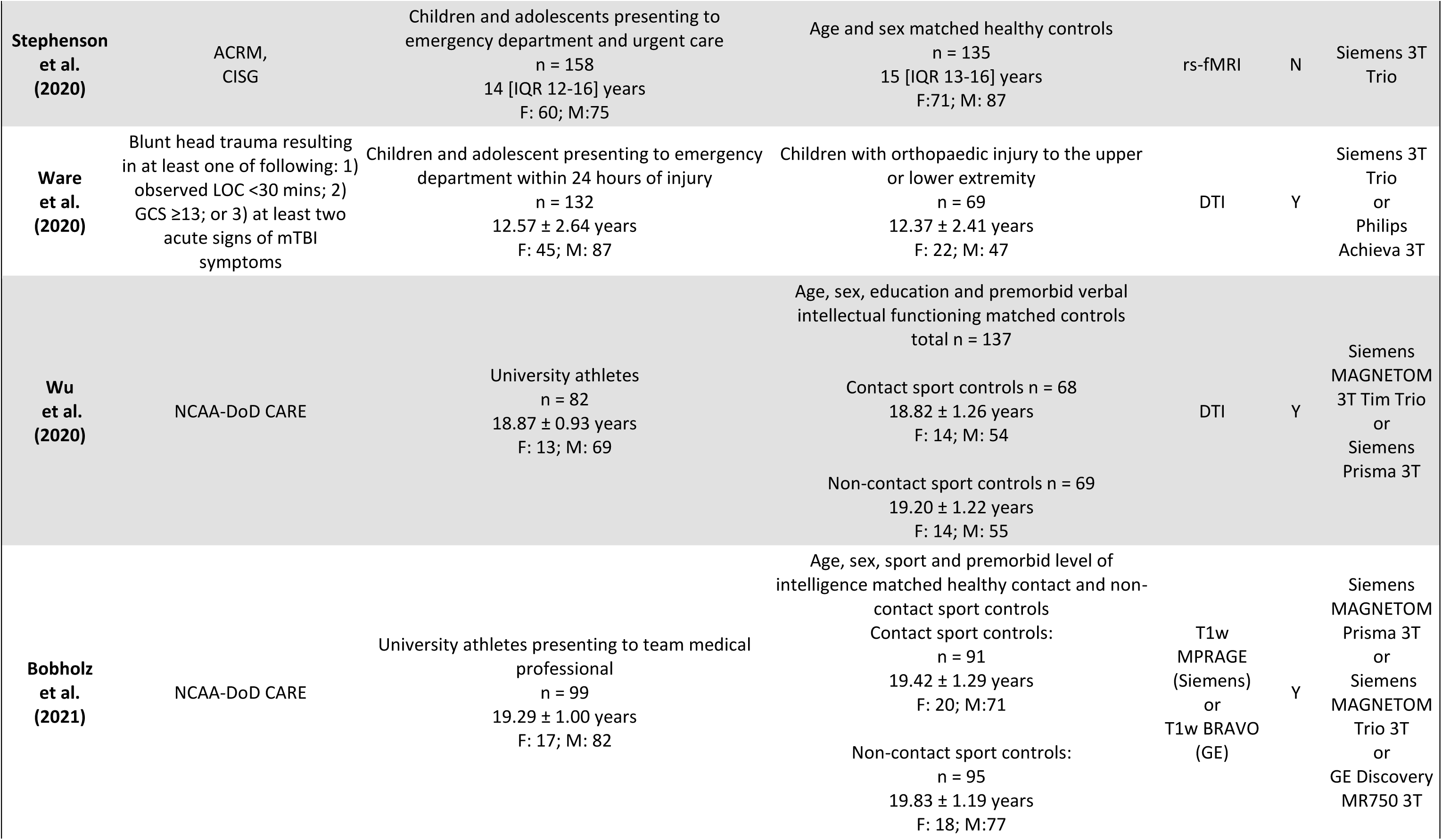
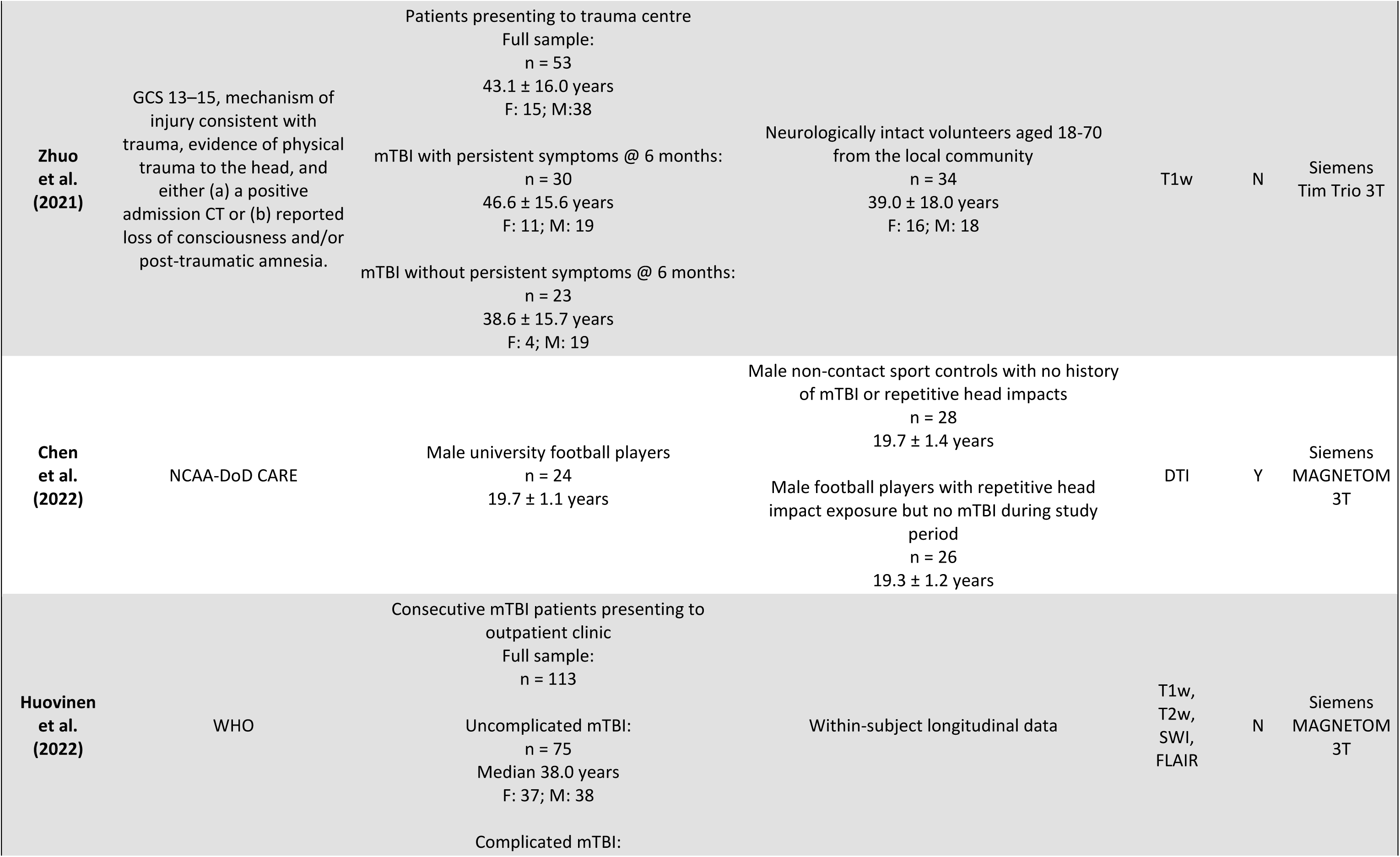
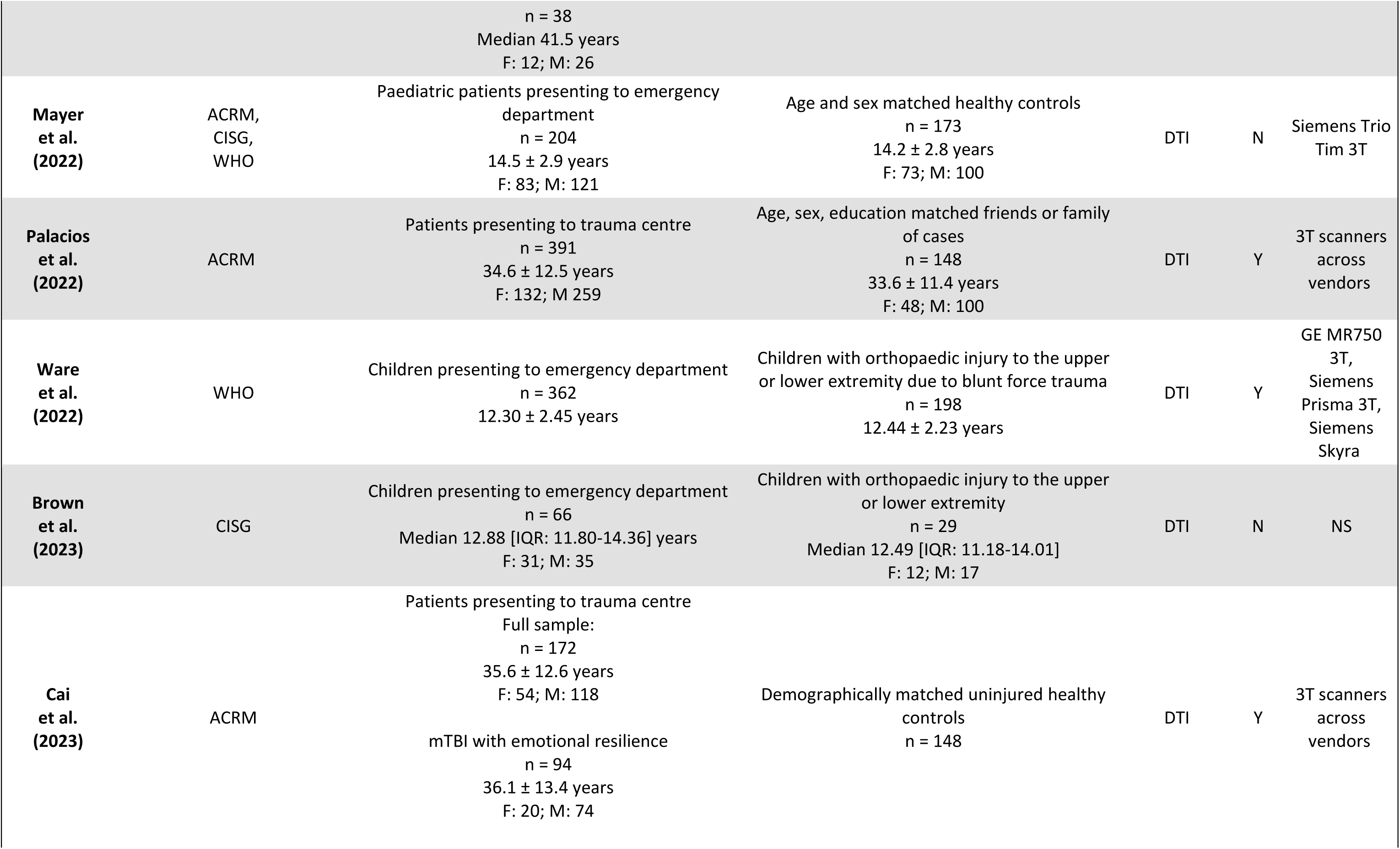
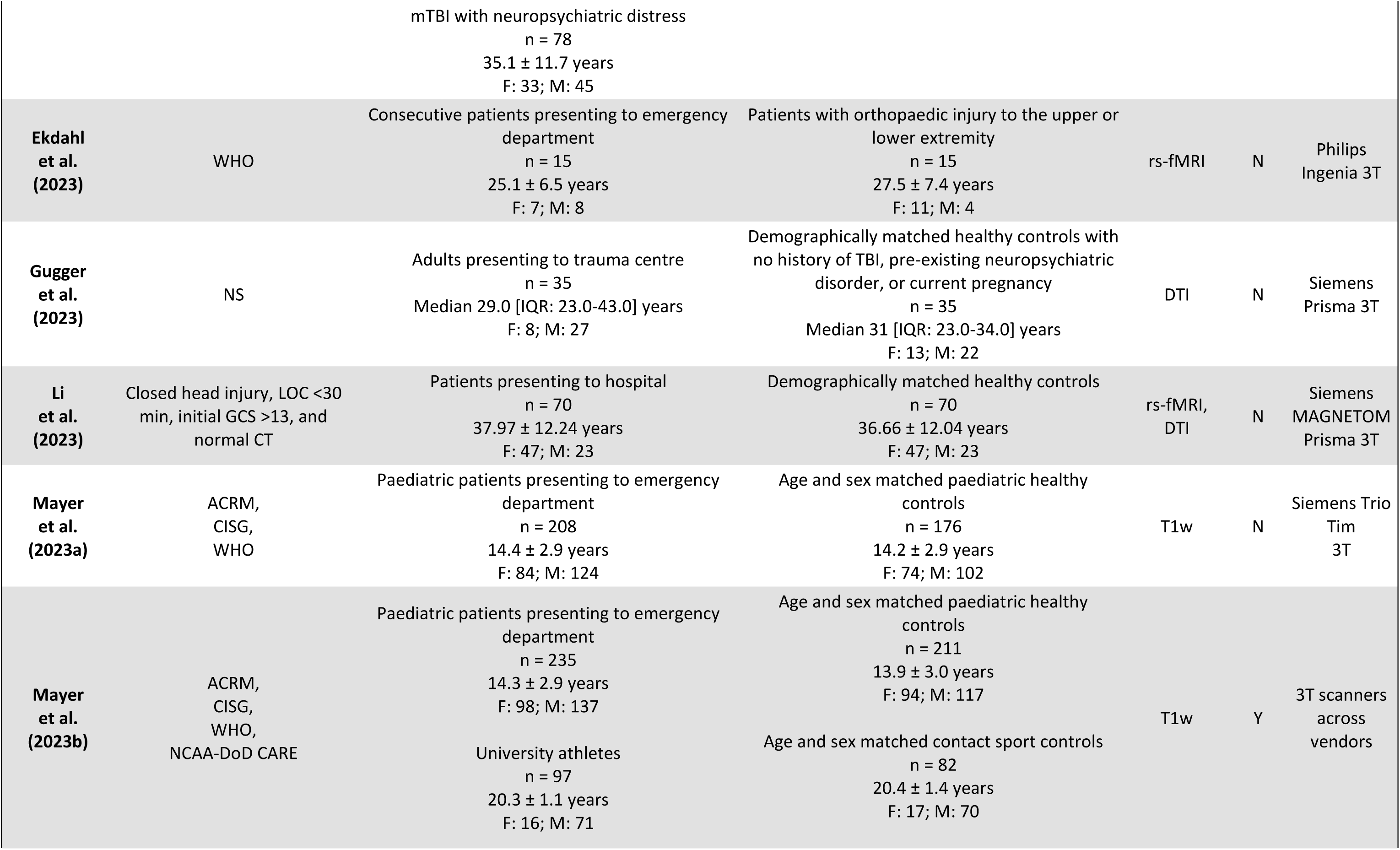
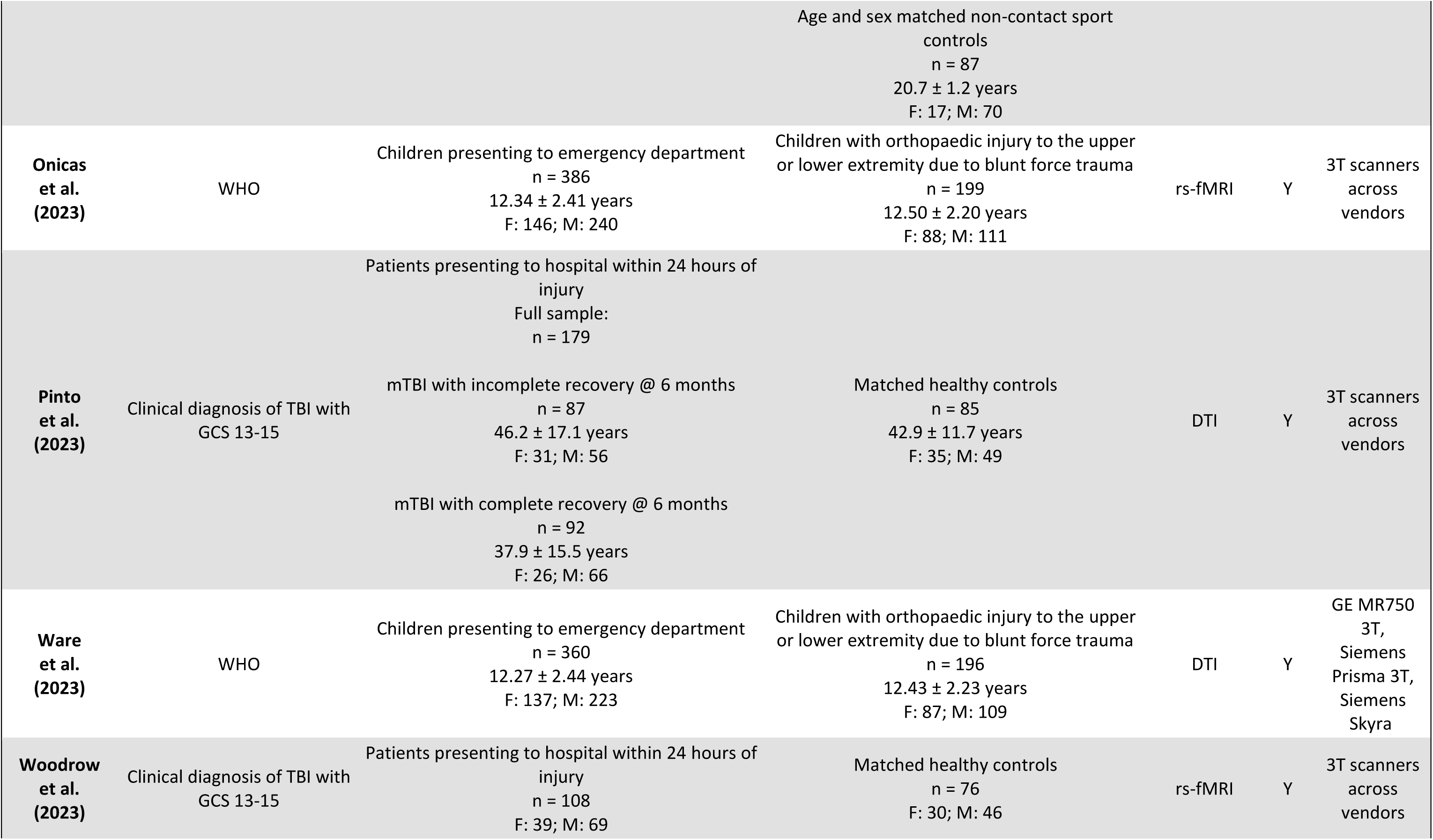
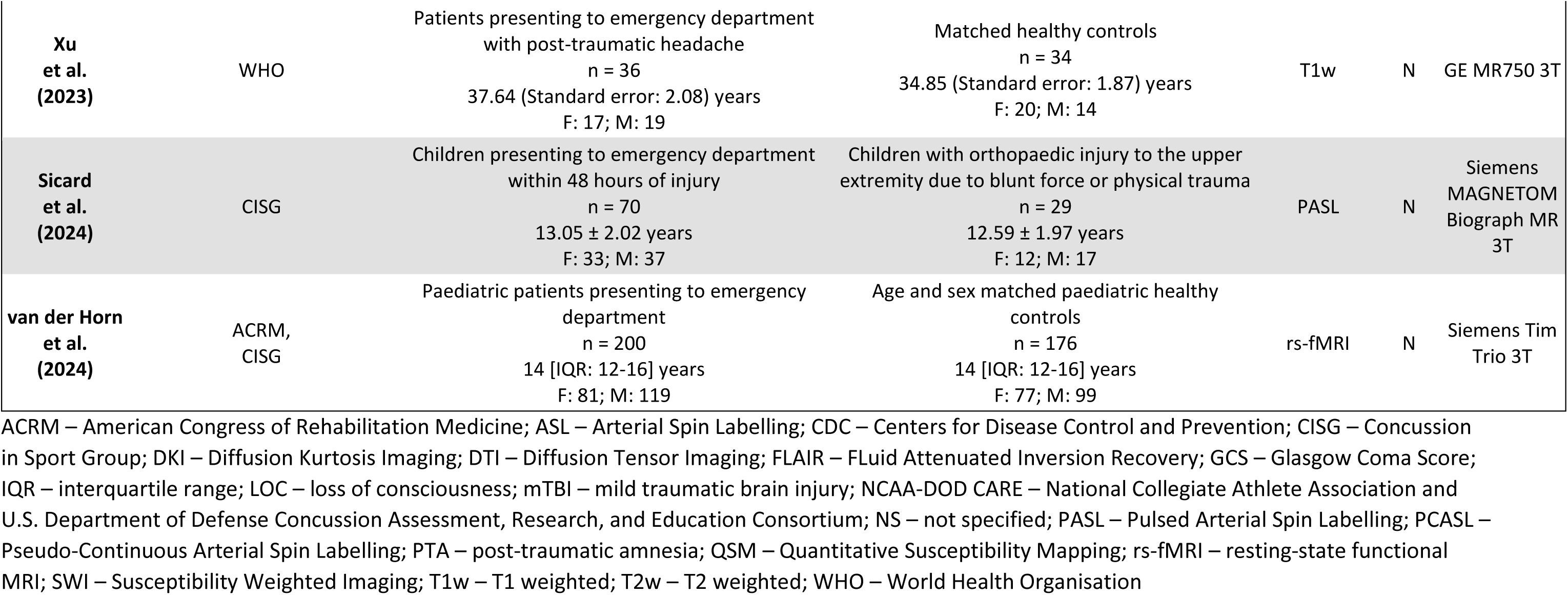
Summary of studies meeting inclusion for review.

All studies adopted an observational cohort design. There were three main cohorts of mTBI patients assessed with 17 studies investigating sport-related mTBI (Henry et al. 2011, Meier et al. 2015a, Meier et al. 2016, Churchill et al. 2017, Manning et al. 2017, Meier et al. 2017, Koch et al. 2018, McCuddy et al. 2018, Murdaugh et al. 2018, Stephens et al. 2018, Churchill et al. 2019a, Churchill et al. 2019b, Wang et al. 2019, Meier et al. 2020, Muftuler et al. 2020, Wu et al. 2020, Bobholz et al. 2021, Chen et al. 2022), 14 studies evaluating paediatric mTBI (Baker et al. 2020, Mayer et al. 2020a, Stephenson et al. 2020, Ware et al. 2020, Fleck et al. 2021, Mayer et al. 2022, Ware et al. 2022, Brown et al. 2023, Mayer et al. 2023a, Mayer et al. 2023b, Onicas et al. 2023, Ware et al. 2023, Sicard et al. 2024, van der Horn et al. 2024) and 20 studies examining consecutive mTBI patients presenting to emergency departments, trauma centres, or outpatient facilities (Yuh et al. 2014, Sours et al. 2015a, Sours et al. 2015b, Dall’Acqua et al. 2017, Palacios et al. 2017, Evans et al. 2018, Studerus-Germann et al. 2018, Madhavan et al. 2019, Yin et al. 2019, D’Souza et al. 2020, Zhuo et al. 2021, Huovinen et al. 2022, Palacios et al. 2022, Cai et al. 2023, Ekdahl et al. 2023, Gugger et al. 2023, Li et al. 2023, Pinto et al. 2023, Woodrow et al. 2023, Xu et al. 2023). Comparators/control cohorts reported across the studies included age and sex-matched healthy controls (Sours et al. 2015a, Sours et al. 2015b, Dall’Acqua et al. 2017, Palacios et al. 2017, Studerus-Germann et al. 2018, Madhavan et al. 2019, Yin et al. 2019, Baker et al. 2020, D’Souza et al. 2020, Mayer et al. 2020a, Stephenson et al. 2020, Wu et al. 2020, Bobholz et al. 2021, Zhuo et al. 2021, Mayer et al. 2022, Palacios et al. 2022, Cai et al. 2023, Gugger et al. 2023, Li et al. 2023, Mayer et al. 2023a, Mayer et al. 2023b, Pinto et al. 2023, Woodrow et al. 2023, Xu et al. 2023, van der Horn et al. 2024), matched healthy contact sport athletes (Meier et al. 2015a, Meier et al. 2016, Manning et al. 2017, Meier et al. 2017, Murdaugh et al. 2018, Churchill et al. 2019b, Wang et al. 2019, Meier et al. 2020, Wu et al. 2020, Bobholz et al. 2021, Chen et al. 2022, Mayer et al. 2023b) matched healthy non-contact sport athletes (Henry et al. 2011, Churchill et al. 2017, Koch et al. 2018, McCuddy et al. 2018, Stephens et al. 2018, Churchill et al. 2019a, Churchill et al. 2019b, Muftuler et al. 2020, Chen et al. 2022, Mayer et al. 2023b), patients with isolated upper/lower extremity orthopaedic injury (Mayer et al. 2020a, Ware et al. 2020, Fleck et al. 2021, Ware et al. 2022, Brown et al. 2023, Ekdahl et al. 2023, Onicas et al. 2023, Ware et al. 2023, Sicard et al. 2024) or within-subject longitudinal data (Yuh et al. 2014, Evans et al. 2018, Huovinen et al. 2022). Approximately half of controls/comparators were assessed at a single timepoint, while the remainder were assessed at multiple timepoints after injury.

Eighteen studies used data from registered research initiatives including: A-CAP (Ware et al. 2020, Ware et al. 2022, Onicas et al. 2023, Ware et al. 2023), TRACK-TBI (Yuh et al. 2014, Palacios et al. 2017, Evans et al. 2018, Palacios et al. 2022, Cai et al. 2023), NCAA-DOD-CARE (Wang et al. 2019, Meier et al. 2020, Wu et al. 2020, Bobholz et al. 2021, Chen et al. 2022), CENTER-TBI (Pinto et al. 2023, Woodrow et al. 2023), and PEDCARE (Brown et al. 2023, Sicard et al. 2024). Seventeen studies analysed data collected across multiple sites (Evans et al. 2018, Madhavan et al. 2019, Wang et al. 2019, Mayer et al. 2020a, Meier et al. 2020, Ware et al. 2020, Wu et al. 2020, Bobholz et al. 2021, Chen et al. 2022, Palacios et al. 2022, Ware et al. 2022, Cai et al. 2023, Mayer et al. 2023b, Onicas et al. 2023, Pinto et al. 2023, Ware et al. 2023, Woodrow et al. 2023).

### 3.2 Clinical outcomes

#### 3.2.1 Symptomology and quality of life

In this review, 17 instruments were used to assess mTBI symptom burden (see Tables 3-6). The most common instruments were: Rivermead Post-Concussion Symptoms Questionnaire (RPQ; 13 studies; (King et al. 1995)); Sports Concussion Assessment Tool (SCAT) symptom questionnaire (2^nd^ edition: 1 study, 3^rd^ edition 11 studies; (2009, 2013)), Post-Concussion Symptom Scale (PCSS; 7 studies; (Lovell and Collins 1998)); Post-Concussion Symptom Inventory (PCSI; 6 studies; (Sady et al. 2014)); and Health and Behaviour Inventory (HBI; 5 studies, (Ayr et al. 2009)). Most studies assessed clinical symptomology on the same day as the neuroimaging session. The remaining studies assessed symptomology within 2-3 days of the scan session. Three studies directly assessed the quality-of-life following mTBI using the Paediatric Quality of Life Inventory (Varni et al. 2001).

**Table 3.** Results from studies using structural and/or Susceptibility Weighted Imaging.

| Authors | MRI sequence(s) | Imaging analysis | Imaging timepoints | Measure of symptoms or functional recovery | What does imaging tell us about symptoms and/or functional recovery? |
| --- | --- | --- | --- | --- | --- |
| **Evans et al. (2018)** | T1w, T2w, FLAIR, SWI | Presence or absence of lesions | 2 weeks P-INJ<br>No control group | HBI symptom scores | Presence of extra-axial (but not intra-axial) lesions predicted *lower* HBI cognitive symptoms @ 3 months P-INJ |
| **Koch et al. (2018)** | QSM | ROI analysis of susceptibility | 24 hours P-INJ, 8 days P-INJ, 6 months P-INJ<br>Controls completed 3 timepoints @ same intervals | SCAT3 symptom scores, days until RTP | No correlation between global or subregional susceptibility and SCAT3 symptom scores @ 24 hours P-INJ<br>Delayed RTP positively associated with greater susceptibility measured @ 24 hours P-INJ within the R cingulum, L inferior fronto-occipital fasciculus, L/R inferior longitudinal fasciculus and L/R superior longitudinal fasciculus ($r(s) = 0.58$ to $0.67$) |
| **Studerus-Germann et al. (2018)** | T1w, T2w, SWI | Presence or absence of microbleeds | 3 days P-INJ, 3 months P-INJ, 1 year P-INJ<br>Single timepoint for controls | PCSS scores | ↑ PCSS total symptom score and fatigue, difficulty concentrating and difficulty remembering scores observed in mTBI participants with microbleeds than those without microbleeds @ 1 year P-INJ<sup>†</sup><br>Number of microbleeds @ 3 days P-INJ in the mTBI group positively associated with greater PCSS total symptom score and PCSS cognitive/emotional items @ 3 months and 1 year P-INJ<sup>†</sup> |
| **Mayer et al. (2020a)** | T1w, T2w, SWI, T2*w, FLAIR | Pathoanatomic features of mTBI based on radiological common data elements | 7 days P-INJ, 3-4 months P-INJ<br>Orthopaedic injury controls completed 2 timepoints @ same intervals | Parent and child PCSI and HBI symptom scores, PedsQL™ scores | No main effect or interaction effect between MRI findings and PCSI or HBI symptom reports @ ~ 1 week or ~3/4 months P-INJ<br>No main effect or interaction effect between MRI findings and PedsQL™ scores @ ~3/4 months P-INJ |
| **Bobholz et al. (2021)** | T1w | Vertex-wise cortical thickness and cortical volume | 24-48 hours P-INJ, Asymptomatic, 7 days post-RTP, 6 months P-INJ<br>Controls completed 4 timepoints @ same intervals | SCAT3 symptom scores, days until asymptomatic | ND in subcortical volumes within the thalamus, hippocampus, or dorsal striatum in mTBI when symptomatic (24-48 hours P-INJ) or asymptomatic |
| **Zhuo et al. (2021)** | T1w | Subcortical volumes | ≤10 days P-INJ, 1 month P-INJ, 6 months P-INJ<br>Single timepoint for controls | GOS-E scores, RPQ symptom scores, presence of persistent symptoms @ 6-months P-INJ | ND subcortical volumes within mTBI without persistent symptoms @ 6 months @ any timepoint<br>↓ thalamic volume within mTBI with persistent symptoms @ 6 months from ≤10 days P-INJ to 6 months P-INJ<br>↓ volume from ≤10 days P-INJ to 6 months P-INJ within the thalamus, hippocampus, amygdala, and putamen associated with ↓ GOS-E ($r(s) = 0.49$ to $0.56$) and ↑ RPQ symptom scores ($r(s) = -0.43$ to $-0.55$) @ 6 months P-INJ |
| **Huovinen et al. (2022)** | T1w, T2w, SWI, FLAIR | Pathoanatomic features of mTBI based on radiological common data elements | 10 days P-INJ<br>No control group | RPQ symptom scores, GOS-E score, time to return to work | ↑ days to full return to work and RPQ symptom scores in complicated mTBI vs uncomplicated mTBI<br>↑ days to full return to work in complicated mTBI with >1 traumatic intracranial lesion vs complicated mTBI with 1 lesion<br>↑ days to full return to work in complicated mTBI with subdural haemorrhages, subarachnoid haemorrhages, or cerebral contusions vs complicated mTBI with other types of lesions |
| **Mayer et al. (2023a)** | T1w | Cortical thickness surface maps and ROI volumes for the bilateral hippocampi and subcortical structures | 7 days P-INJ, 4 months P-INJ<br>Controls completed 2 timepoints @ same intervals | PCSI symptom scores and 5P risk score<br>Persistent symptom | ND cortical thickness symptomatic mTBI or asymptomatic mTBI vs controls @ 7 days or 4 months P-INJ<br>ND hippocampal volume symptomatic mTBI vs asymptomatic mTBI @ 7 days or 4 months P-INJ |
|  |  |  |  | status @ 4 months P-INJ | A random forest model using 5P risk score, clinical symptom data and cortical thickness of the rostral middle frontal gyrus and volume of the hippocampi classified mTBI vs control more accurately than a model using only 5P risk score and clinical symptom data (AUC = 0.815 vs 0.731; balance accuracy = 0.746 vs 0.707, sensitivity = 0.732 vs 0.729, specificity = 0.761 vs 0.686) |
| **Mayer et al. (2023b)** | T1w | T1w images were used to estimate brain age and calculate predicted age difference | Paediatric cohort: 7 days P-INJ, 4 months P-INJ<br>University cohort: ≤2 days P-INJ, asymptomatic, 7 days post-RTP<br>Controls completed 2-3 timepoints @ same interval | Paediatric cohort: PCSI symptom scores and 5P risk score<br>University cohort: SCAT3 symptom score | Negative association between 5P risk score and predicted age difference in paediatric cohort @ 7 days P-INJ<br>No association between predicted age difference and SCAT3 symptom scores in university-aged mTBI cohort @ ≤2 days P-INJ, asymptomatic, or 7 days post-RTP timepoints |
| **Xu et al. (2023)** | T1w | Cortical thickness and cortical surface area | ≤7 days P-INJ, 1 month P-INJ<br>Controls completed 2 timepoints @ same intervals | Head Impact Test symptom score | ↑ % change in left anterior cingulate cortex ($r = 0.33$) and left insula ($r = 0.41$) from ≤7 days to 1 month P-INJ positively associated with ↑ % change in Head Impact Test symptom scores |

† - no multiple comparisons correction reported; AUC – area under the curve; FLAIR – FLuid Attenuated Inversion Recovery; GOS-E – Glasgow Outcome Scale Extended; HBI – Health and Behaviour Inventory; mTBI – mild traumatic brain injury; ND – no difference; PCSI – Post Concussion Symptom Inventory; PCSS – Post Concussion Symptom Scale; PedsQL™ - Paediatric Quality of Life Inventory; P-INJ – post-injury; QSM – Quantitative Susceptibility Mapping; ROI – region-of-interest; RTP – return to play; RPQ – Rivermead Post Concussion Symptoms Questionnaire; SCAT – Sport Concussion Assessment Tool; SWI – susceptibility weighted imaging; T1w – T1 weighted; T2w – T2 weighted

Six studies used symptom data for subgroup/phenotype analysis within the mTBI sample. Relationships between cognitive, emotional, and somatic symptom subscales and neuroimaging findings were evaluated in three studies. The three remaining studies compared neuroimaging results for mTBI participants with low versus high symptom burden, participants with/without cognitive intolerance, and mTBI participants demonstrating neuropsychiatric distress versus those with emotional resilience.

#### 3.2.2 Definitions of recovery

There was significant heterogeneity in the operational definitions of mTBI recovery status across studies in this review (see Tables 3-6). The number of days until becoming asymptomatic, returning to play (RTP), or returning to work was used as a measure of time until functional recovery in seven studies. Recovery status was measured with the Glasgow Outcome Scale – Extended (GOS-E; (Wilson et al. 2021) either at 3-4 months post-injury (P-INJ) in two studies, or at six months P-INJ in a further seven studies. Fifteen studies differentiated recovered mTBI participants (those with resolved symptoms) from participants with persistent symptoms using available symptom data. However, the criteria used for classifying recovery status varied across studies. Six studies determined persistent symptom status at six months P-INJ based on features consistent with ICD-10 criteria (World Health Organization 2004). Two studies used simple thresholds to define persistent symptom status. The first study based persistent symptom status on the symptom count being >1 at six weeks P-INJ, while the second study used a total PCSS symptom score of >8 for females and >6 for males. Seven studies implemented standardized change metrics (*z*-score based) to establish persistent status. Four studies compared post-injury symptom scores to premorbid symptom reports, and the final three compared against ratings from the control group (Mayer et al. 2020b).

### 3.3 Imaging findings

This review encompasses a range of neuroimaging techniques used to investigate mTBI symptomology and functional recovery, each offering distinct insights into brain structure and function. The imaging findings are summarized in four themes: 1) structural and SWI imaging; 2) diffusion-weighted imaging approaches; 3) blood-oxygenation level dependent (BOLD) imaging; and 4) ASL imaging. One study included Magnetic Resonance Spectroscopy but did not report any findings within the scope of this review. Figure 2 illustrates the MRI modalities used in the studies covered by this review, categorizing them according to each of the four themes and highlighting their specific applications.

**Figure 2.**
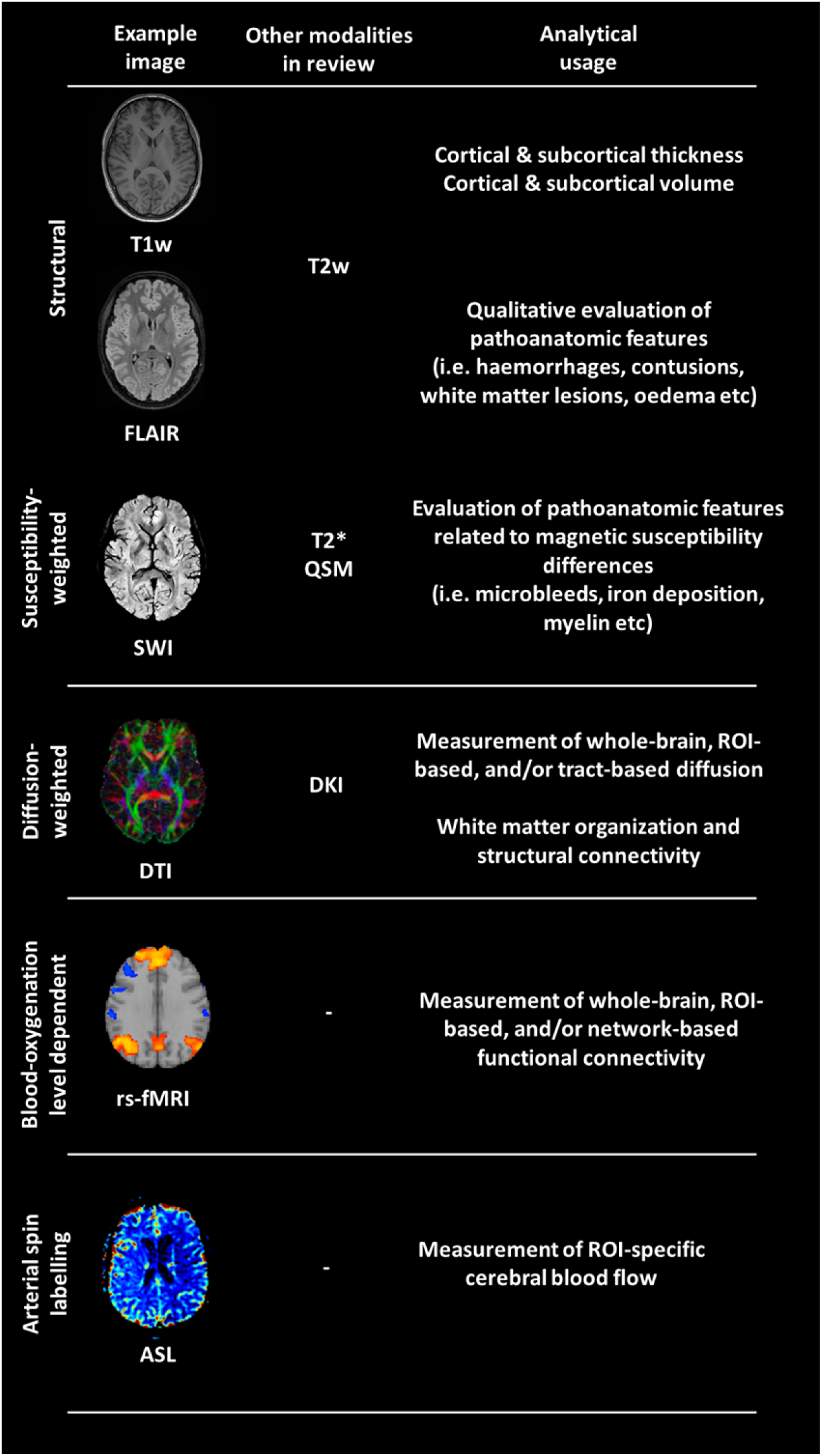
Summary of MRI modalities used across studies. T1w – T1-weighted; T2w – T2-weighted; FLAIR – FLuid Attenuated Inversion Recovery; SWI – Susceptibility Weighted Imaging; QSM – Quantitative Susceptibility Mapping; DTI – Diffusion Tensor Imaging; DKI – Diffusion Kurtosis Imaging; rs-fMRI – Resting-state Functional MRI; ASL – Arterial Spin Labelling.

#### 3.3.1 Structural and susceptibility-weighted imaging and functional recovery outcomes

Ten studies reported findings exploring relationships between widely used conventional structural imaging (i.e., being commonly used in a hospital setting) and SWI methods and measures of mTBI symptoms and/or functional recovery (Table 3). The most common sequence used was T1w (9/10 studies), followed by SWI (4/10 studies), T2w (4/10 studies), FLAIR (3/10 studies), with QSM and T2* sequences each used in a single study.

Four studies combined three or more of these sequences to evaluate pathoanatomic features of mTBI (i.e. microbleeds, haemorrhages, contusions, etc.) and their association with clinical outcomes (Evans et al. 2018, Studerus-Germann et al. 2018, Mayer et al. 2020a, Huovinen et al. 2022). In one study (total n = 50), the number of microbleeds at 3 days P-INJ was positively associated with higher symptom scores (PCSS) at three months and one year P-INJ (Studerus-Germann et al. 2018). Similarly, higher symptom scores were observed in mTBI participants with pathoanatomic features than those without (total n = 113; (Huovinen et al. 2022)). Participants with >1 lesion took longer to return to work than those with a single lesion and subdural/subarachnoid haemorrhages and cerebral contusions were associated with a longer time to return to work (Huovinen et al. 2022). In contrast, one study (total n = 45) found that extra-axial lesions predicted lower symptoms (measured by HBI) at three months P-INJ (Evans et al. 2018), while another study (total n = 464) found no association between pathoanatomic MRI findings and symptom reports or QOL at one week and 3-4 months P-INJ (Mayer et al. 2020a).

Relationships between mTBI symptoms and functional recovery and QSM findings were evaluated in one study (total n = 56; (Koch et al. 2018)). The authors reported no relationship between global or regional susceptibility and SCAT3 symptom scores at 24 hours P-INJ, however, susceptibility at this timepoint within subregions (See Table 3) demonstrated moderate to strong positive correlations with delayed return to play (RTP) following sport-related mTBI.

The remaining five studies derived quantitative measures from T1w images and correlated these measures with clinical outcomes. Four of five studies calculated cortical thickness and cortical/sub-cortical volumes from T1w images (Bobholz et al. 2021, Zhuo et al. 2021, Mayer et al. 2023a, Xu et al. 2023). The final study used T1w to estimate brain age and calculate the predicted age difference to see if this related to mTBI recovery outcomes (Mayer et al. 2023b). Two studies (total n = 285 and 384) found no differences in cortical thickness and/or cortical/sub-cortical volumes based on symptomatic status (Bobholz et al. 2021, Mayer et al. 2023a). A third study (total n = 87) reported no within-group differences in subcortical volumes in mTBI patients with resolved symptoms spanning 10 days, one month, and six months P-INJ (Zhuo et al. 2021). However, reduced thalamic volume was detected from 10 days to six months P-INJ in patients with persistent symptoms. Reduced volume from 10 days to 6 months P-INJ within the thalamus, hippocampus, amygdala, and putamen all demonstrated moderate positive associations with symptom scores (measured by RPQ) and moderate negative associations with recovery outcome (measured by GOS-E) at six months P-INJ (Zhuo et al. 2021). Another study (total n = 70) observed greater changes in the left anterior cingulate cortex and left insula from seven days to one month P-INJ demonstrated a weak-moderate positive relationship with change in symptom scores (measured by Head Impact Test) over the same interval (Xu et al. 2023). Finally, a negative association between recovery risk score (measured by 5P risk score) and predicted age difference was reported in a large study (total n = 712) for children with mTBI at seven days P-INJ (Mayer et al. 2023b). However, no association between predicted age difference and symptom scores (measured by SCAT) were observed in university-aged acute mTBI patients or when asymptomatic (Mayer et al. 2023b).

#### 3.3.2 Diffusion-weighted imaging approaches and functional recovery outcomes

Diffusion-weighted imaging sequences were the most common imaging modality in this review, utilized in 26 studies (Table 4). Twenty-five studies used Diffusion Tensor Imaging (DTI), with one study also including neurite orientation dispersion and density imaging (NODDI) and another also including tract density imaging, and the last study using Diffusion Kurtosis Imaging (DKI) (Henry et al. 2011, Yuh et al. 2014, Meier et al. 2016, Churchill et al. 2017, Dall’Acqua et al. 2017, Manning et al. 2017, Murdaugh et al. 2018, Churchill et al. 2019a, Churchill et al. 2019b, Yin et al. 2019, Baker et al. 2020, Muftuler et al. 2020, Ware et al. 2020, Wu et al. 2020, Fleck et al. 2021, Chen et al. 2022, Mayer et al. 2022, Palacios et al. 2022, Ware et al. 2022, Brown et al. 2023, Cai et al. 2023, Gugger et al. 2023, Li et al. 2023, Pinto et al. 2023, Ware et al. 2023).

**Table 4.**
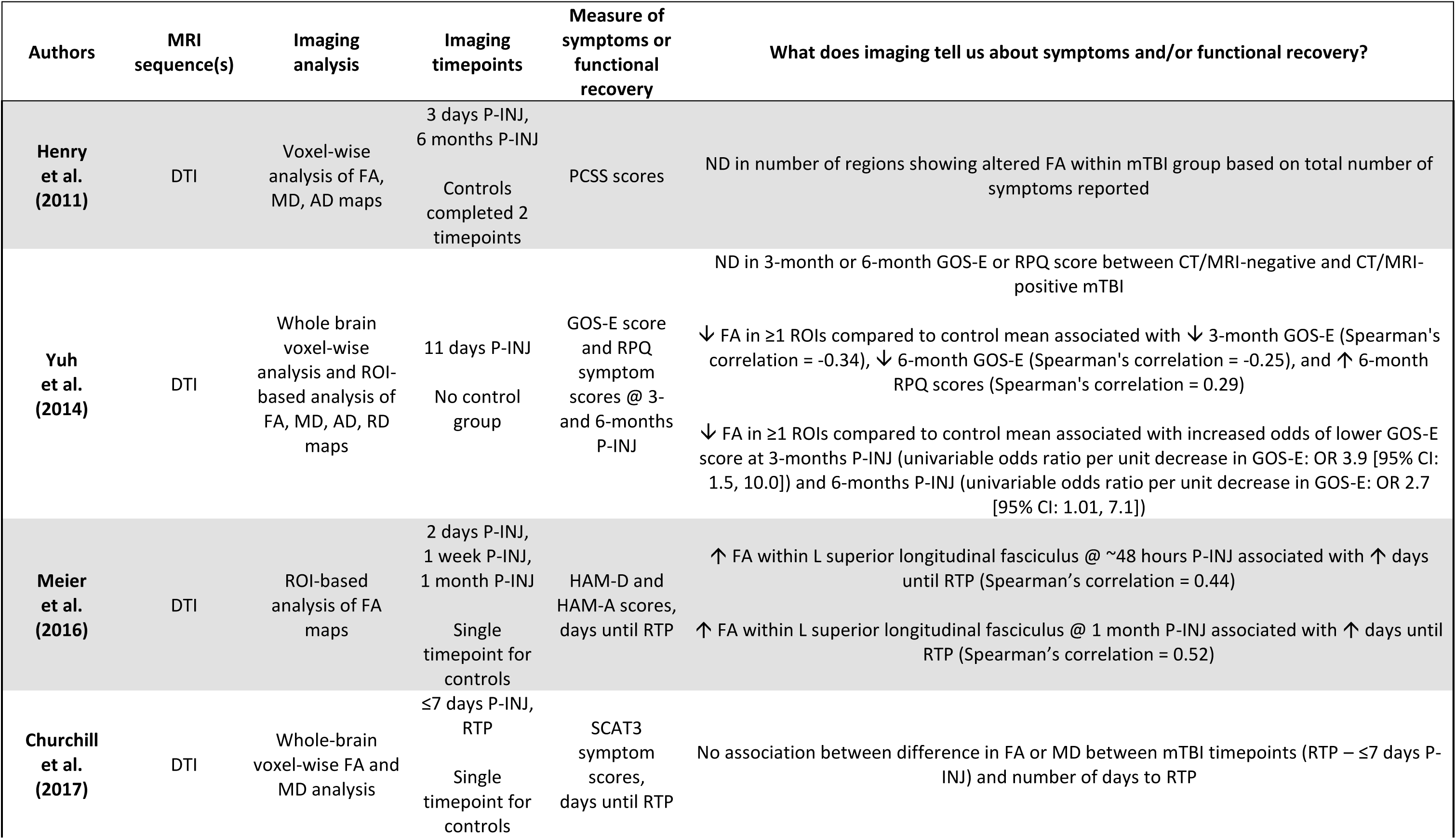
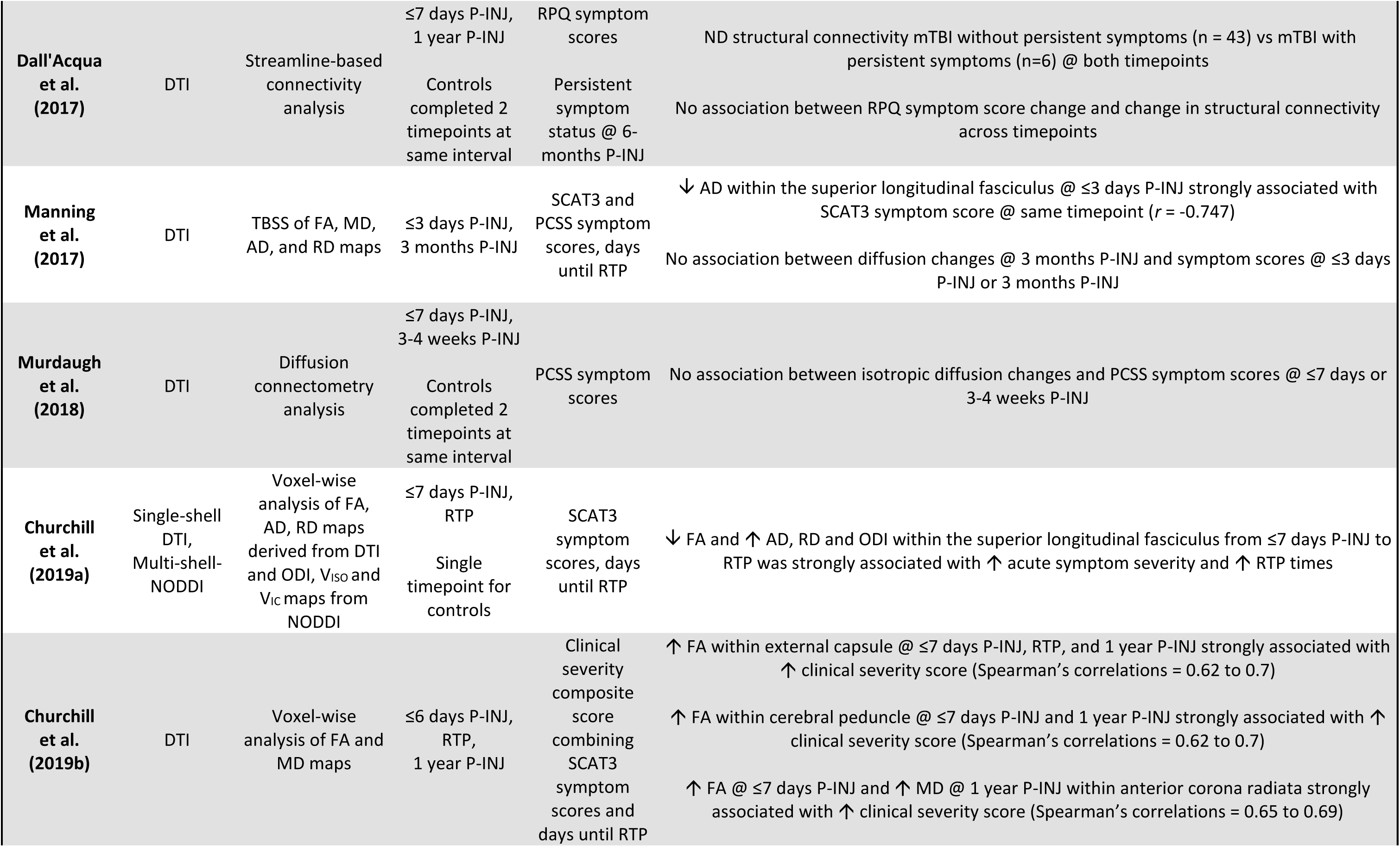
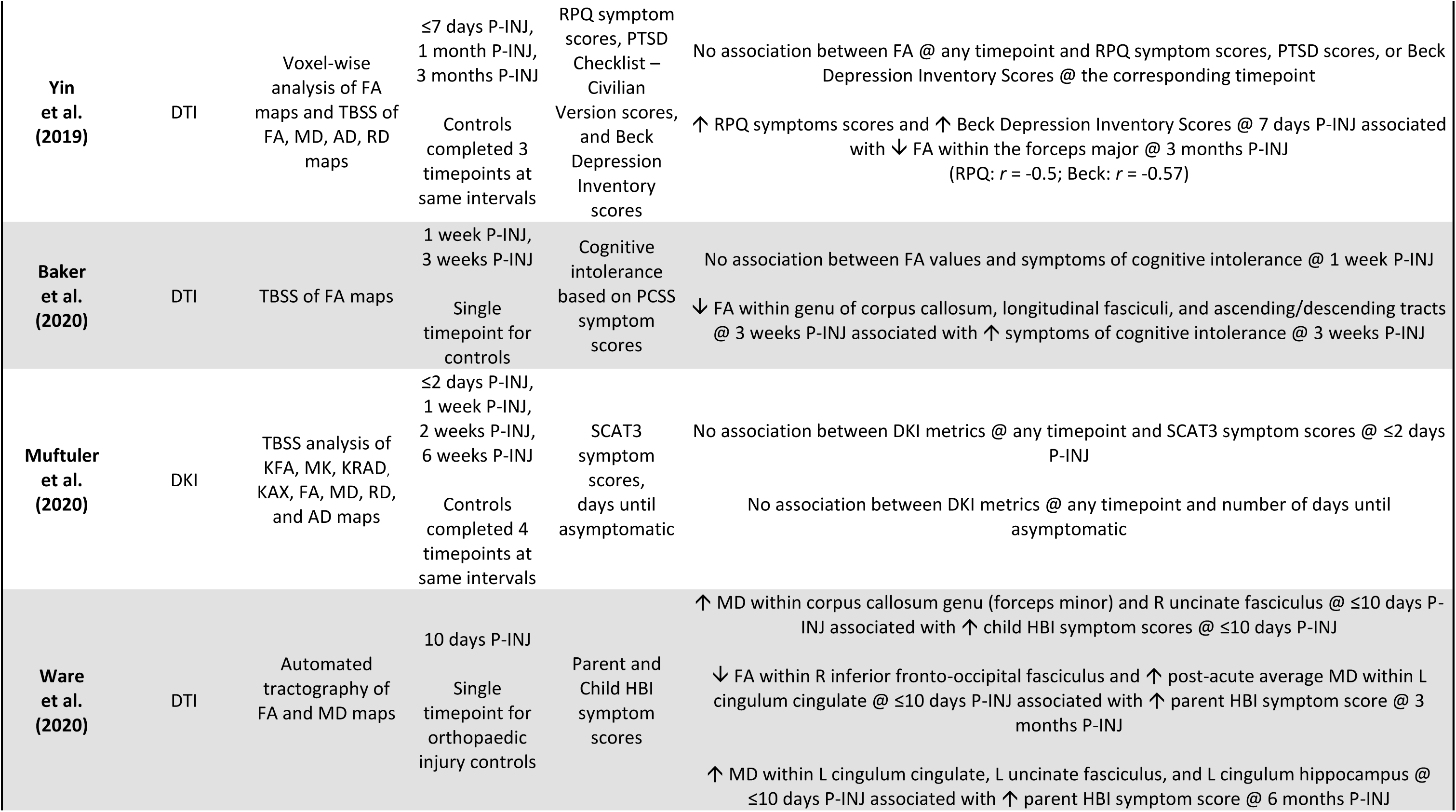
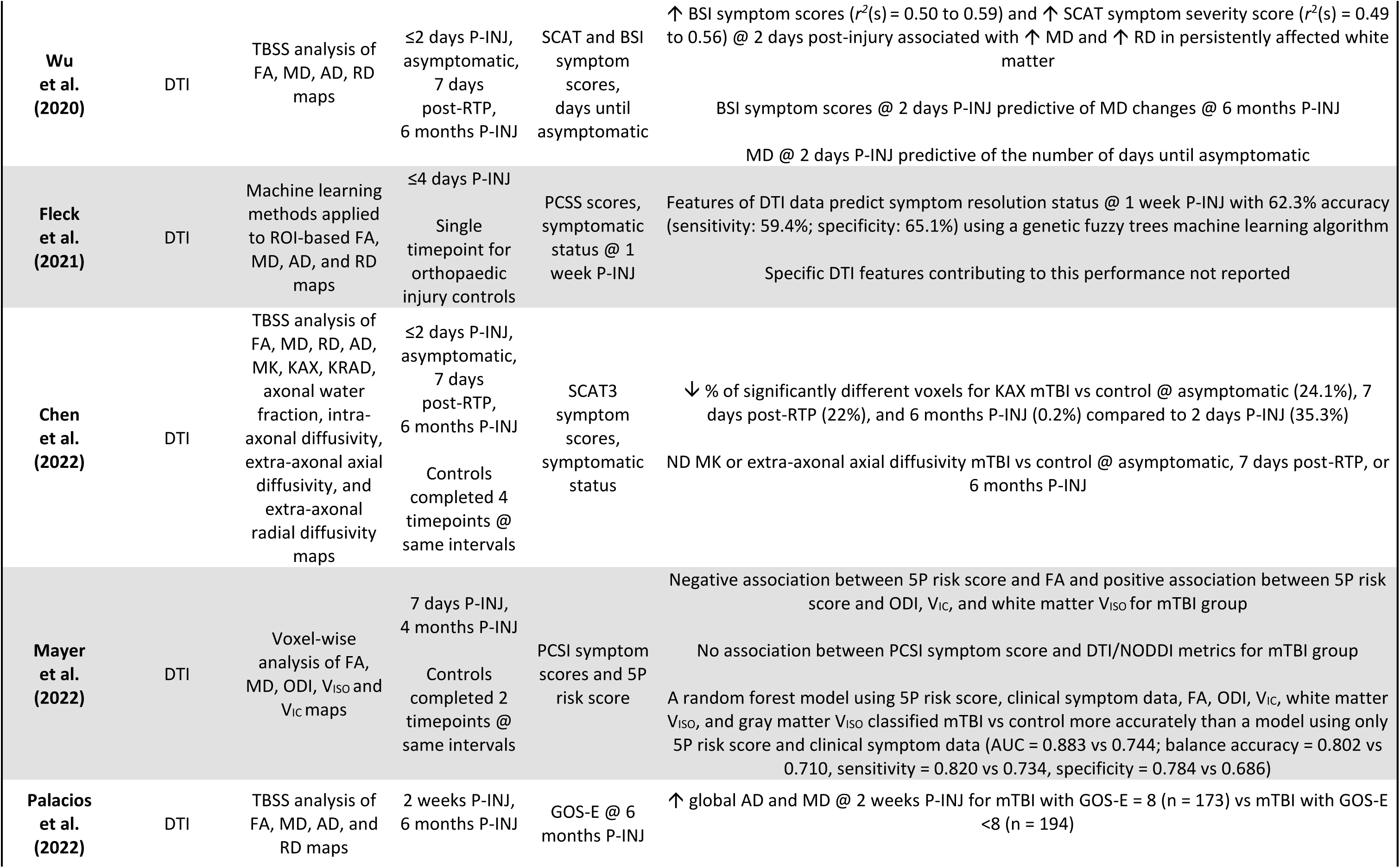
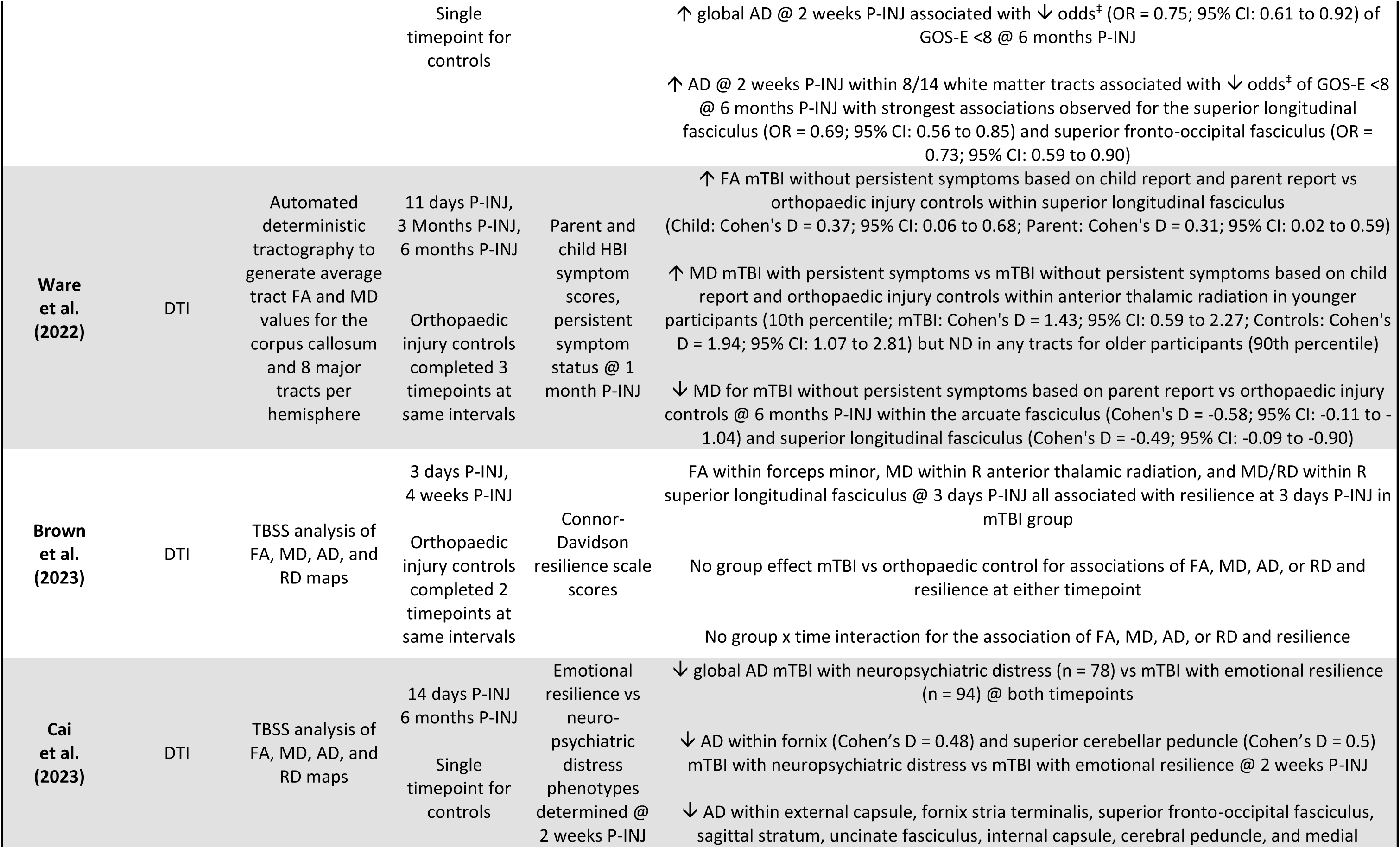
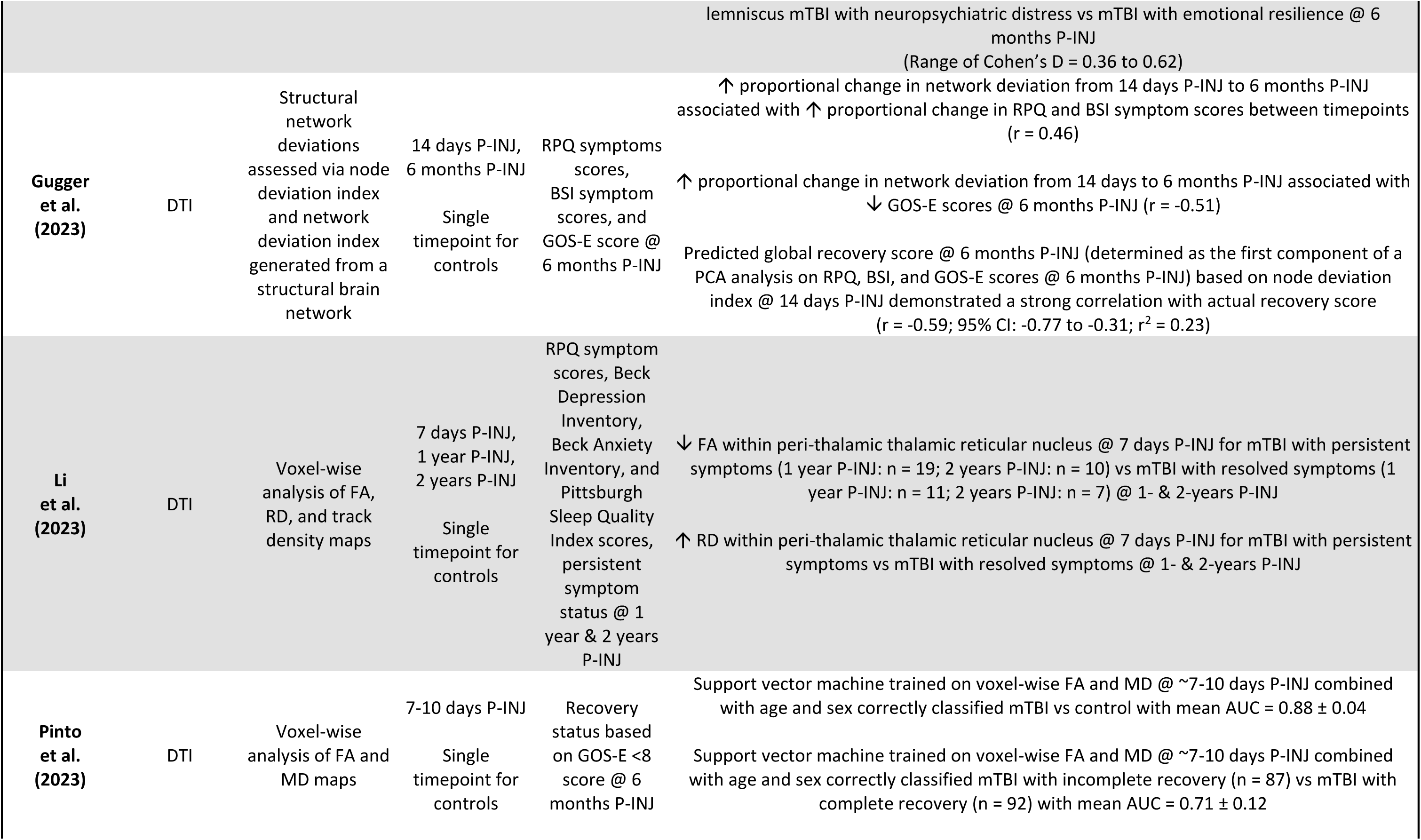
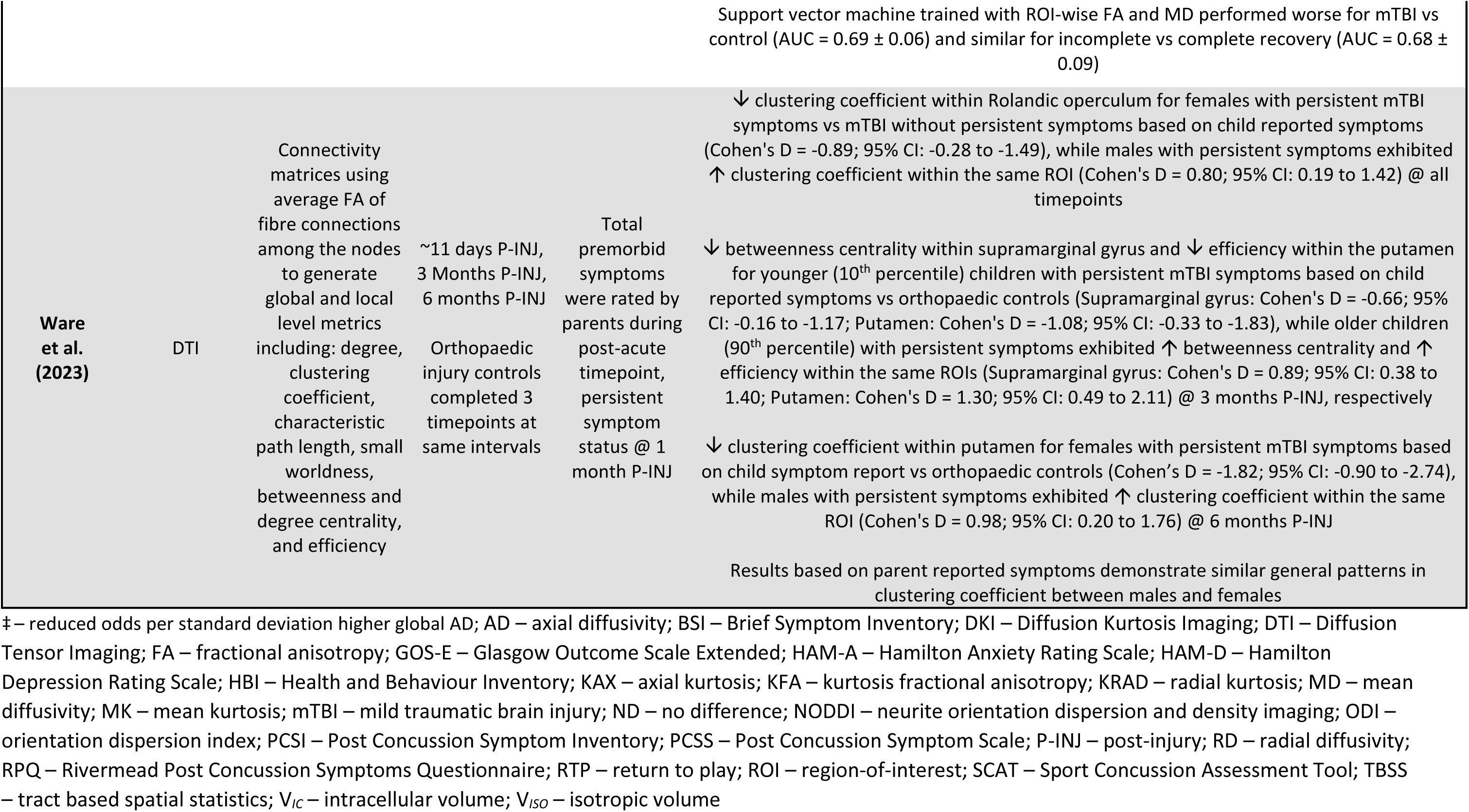
Results from studies using Diffusion-Weighted Imaging approaches.

In eight studies, tract-based spatial statistics (TBSS; (Smith et al. 2006)) were utilised to analyse maps of fractional anisotropy (FA), mean diffusivity (MD), axial diffusivity (AD), and/or radial diffusivity (RD) or DKI-derived maps (Yin et al. 2019, Baker et al. 2020, Muftuler et al. 2020, Wu et al. 2020, Chen et al. 2022, Palacios et al. 2022, Brown et al. 2023, Cai et al. 2023). Two studies (total n = 39 and 178) reported no association between TBSS results from two- or seven-days P-INJ and symptom scores (measured by PCSS or SCAT) at each respective timepoint (Baker et al. 2020, Muftuler et al. 2020). A small study (total n = 43) described a strong association between greater acute symptom scores (measured by SCAT3) and acutely reduced AD within the superior longitudinal fasciculus (Manning et al. 2017). Baker et al. (2020; total n = 39) described reduced FA in multiple tracts were associated with increased symptoms of cognitive intolerance (measured by PCSS) at three weeks P-INJ. Chen et al. (2022; total n = 78) observed no differences in mean kurtosis or extra-axonal AD between the mTBI group when asymptomatic or at six months P-INJ and controls. However, the authors reported that the initial differences in the percentage of voxels with significantly different axial kurtosis between acute symptomatic mTBI patients and controls progressively decreased over time; from 35% at two days P-INJ, to 25% when asymptomatic and <1% at six months P-INJ (Chen et al. 2022). One study (total n = 320) assessed the relationship between self-reported resilience within the mTBI group and TBSS results and found associations between tract-based FA, MD, and RD at 3 days P-INJ. Moderate reductions in AD were observed in multiple tracts at two weeks and six months P-INJ between mTBI participants demonstrating a neuropsychiatric distressed phenotype versus those demonstrating emotional resilience (Cai et al. 2023). Two studies found that acute TBSS findings were predictive of mTBI recovery outcomes. The first study (total n = 219) observed that MD at two days P-INJ was predictive of the number of days until asymptomatic (Wu et al. 2020); while the second study (total n = 539) reported that increased AD at two weeks P-INJ reduced the odds of incomplete recovery (measured by GOS-E) at six months P-INJ (Palacios et al. 2022). A further pair of studies found that worse acute symptoms (measured by SCAT, BSI, RPQ, or Beck Depression Inventory) were moderately associated with TBSS results in persistently affected white matter. Specifically, worse symptoms at two days P-INJ were moderately associated with increased MD and RD at 6 months P-INJ with acute symptoms predictive of MD changes at the timepoint (total n = 219; (Wu et al. 2020)). Worse symptoms at seven days P-INJ were also moderately associated with reduced FA at three months P-INJ (total n = 64; (Yin et al. 2019)).

Nine studies employed voxel-wise analysis of one or more FA, MD, AD, RD maps (Henry et al. 2011, Yuh et al. 2014, Churchill et al. 2017, Churchill et al. 2019a, Churchill et al. 2019b, Yin et al. 2019, Mayer et al. 2022, Li et al. 2023, Pinto et al. 2023). Similar to TBSS findings, three studies (total n = 24, 64, and 377) found no associations between symptom reports (measured by PCSS, PCSI, RPQ, PTSD, or Beck Depression Inventory) and voxel-wise DTI/NODDI metrics collected at the same timepoint (Henry et al. 2011, Yin et al. 2019, Mayer et al. 2022). Churchill et al. (2017; total n = 54) reported no association between the number of days to RTP following sport-related mTBI and differences in voxel-wise FA or MD between the acute and RTP imaging timepoints. In a subsequent study (total n = 66), the authors observed reduced FA and increased AD, RD, and orientation dispersion index (ODI) from the acute imaging timepoint to the RTP imaging timepoint were associated with greater acute symptom severity (measured by SCAT) and greater RTP times (Churchill et al. 2019a). In a third study, Churchill et al. (2019b; total n = 146) found that acutely increased FA within ventral white matter regions was strongly associated with greater acute symptom severity (measured by SCAT3) and longer RTP. The authors also found a positive association between FA and MD at RTP and acute symptom severity and RTP. In one study (total n = 377), acute prolonged recovery risk (measured by 5P risk score) was negatively associated with acute voxel-wise FA and positively associated with acute voxel-wise ODI, intracellular volume (V*_IC_*), and isotropic volume (V*_ISO_*) (Mayer et al. 2022). Another study used voxel-wise DTI measures to predict recovery, reporting moderate classification accuracy of mTBI patients (total n = 264) with incomplete recovery at 6 months P-INJ (measured by GOS-E) using a support vector machine trained on participant age, sex, and acute voxel-wise FA and MD maps (Pinto et al. 2023). Finally, one study used voxel-wise analysis of FA, RD, and tract-density maps (total n = 140; (Li et al. 2023)). They observed reduced FA and increased RD within the peri-thalamic reticular nucleus at 7 days post-injury in mTBI patients with persistent symptoms at 1-2 years P-INJ compared to mTBI patients with resolved symptoms (Li et al. 2023).

Three studies utilised an ROI-based approach to examine specific white matter tracts, either with or without voxel-wise analysis. In one study, the utility of acute voxel-wise or ROI-based FA and/or MD maps to predict incomplete recovery at 6 months P-INJ was assessed. Acutely decreased FA in ≥1 region-of-interest (ROI) compared to the control mean were associated with increased odds of incomplete recovery at 6 months P-INJ (measured by GOS-E) and were weakly associated with increased symptom scores (measured by RPQ) at the same timepoint (total n = 116; (Yuh et al. 2014)). In another study (total n = 86; (Meier et al. 2016)), regions-of-interest corresponding to key white matter tracts were delineated, and increased FA within the superior longitudinal fasciculus at 48 hours and one month after injury showed a moderate positive correlation with days until RTP. Another study utilised ROI-based inputs to train a machine learning classifier to predict symptom recovery from DTI-based measures (FA, MD, AD, and RD). Fleck et al. (2021; n = 43) reported modest classification accuracy (62.3%) of asymptomatic status at one week P-INJ based on a genetic fuzzy trees machine learning algorithm trained on features of DTI data acquired within four days P-INJ.

Two studies used automated tractography and along-tract fibre quantification to explore DTI data (Ware et al. 2020, Ware et al. 2022). The first study (total n = 201) reported positive associations between increased tract-based MD at 10 days P-INJ and increased symptom scores (measured by HBI) at 10 days, three months, and six months P-INJ. Decreased tract-based FA at 10 days P-INJ was also negatively associated with symptoms at three months P-INJ (Ware et al. 2020). In a follow-up study (total n = 560), the authors observed small increases in tract-based FA for children without persistent mTBI symptoms compared to orthopaedic injury controls at 10 days, three months, and 6 months P-INJ (Ware et al. 2022). Moderate decreases in tract-based MD were reported for children without persistent symptoms compared to controls at 6 months P-INJ. Lastly, younger children, but not older children, with persistent mTBI symptoms showed large increases in tract-based FA compared to children without persistent symptoms and controls at all timepoints (Ware et al. 2022).

Structural connectivity analysis was performed on DTI data in four studies (Dall’Acqua et al. 2017, Murdaugh et al. 2018, Gugger et al. 2023, Ware et al. 2023). One small study (total n = 28) reported no association between isotropic diffusion changes and symptom scores (measured by PCSS) at seven days post-injury or one-month post-injury, respectively (Murdaugh et al. 2018). Another study (total n = 98) observed no association between the change in symptom scores (measured by RPQ) and change in structural connectivity from seven days to one year P-INJ and no differences in structural connectivity between mTBI patients with/without persistent symptoms at either timepoint (Dall’Acqua et al. 2017). In contrast, Gugger et al. (2023; total n = 70) found that increased proportional change in network deviation from 14 days to six months P-INJ was moderately associated with increased proportional change in symptom scores (measured by RPQ and BSI) and decreased proportional change in functional recovery (measured by GOS-E) over the same interval. This study also observed that the node deviation (node is a topological representation of a brain region) index at 14 days P-INJ demonstrated a moderate-strong correlation with a principal component analysis derived clinical recovery score (Gugger et al. 2023). The final study (total n = 556) reported large sex- and age-based differences in local structural connectivity metrics between children with persistent mTBI symptoms versus those without persistent symptoms and orthopaedic injury controls (all effect sizes >±0.66). Female children with persistent symptoms at 1 year P-INJ demonstrated a reduced clustering coefficient within multiple ROIs compared to females without symptoms. In contrast, males with persistent symptoms showed higher clustering coefficients at 10 days, three months, and six months P-INJ (Ware et al. 2023). Younger children with persistent symptoms at 1 year P-INJ had decreased betweenness centrality and efficiency within a single ROI compared to orthopaedic injury controls. In contrast, older children with persistent symptoms increased in these metrics relative to controls (Ware et al. 2023).

#### 3.3.3 Blood oxygenation level dependent imaging and functional recovery outcomes

In this review, resting-state functional MRI (rs-fMRI) was the second most common imaging modality (Table 5) used in 18 studies (Sours et al. 2015a, Churchill et al. 2017, Dall’Acqua et al. 2017, Manning et al. 2017, Meier et al. 2017, Palacios et al. 2017, McCuddy et al. 2018, Murdaugh et al. 2018, Churchill et al. 2019b, Madhavan et al. 2019, D’Souza et al. 2020, Meier et al. 2020, Stephenson et al. 2020, Ekdahl et al. 2023, Li et al. 2023, Onicas et al. 2023, Woodrow et al. 2023, van der Horn et al. 2024). No studies explored utility of task-based fMRI to understand mTBI symptomology or functional recovery outcomes.

**Table 5.**
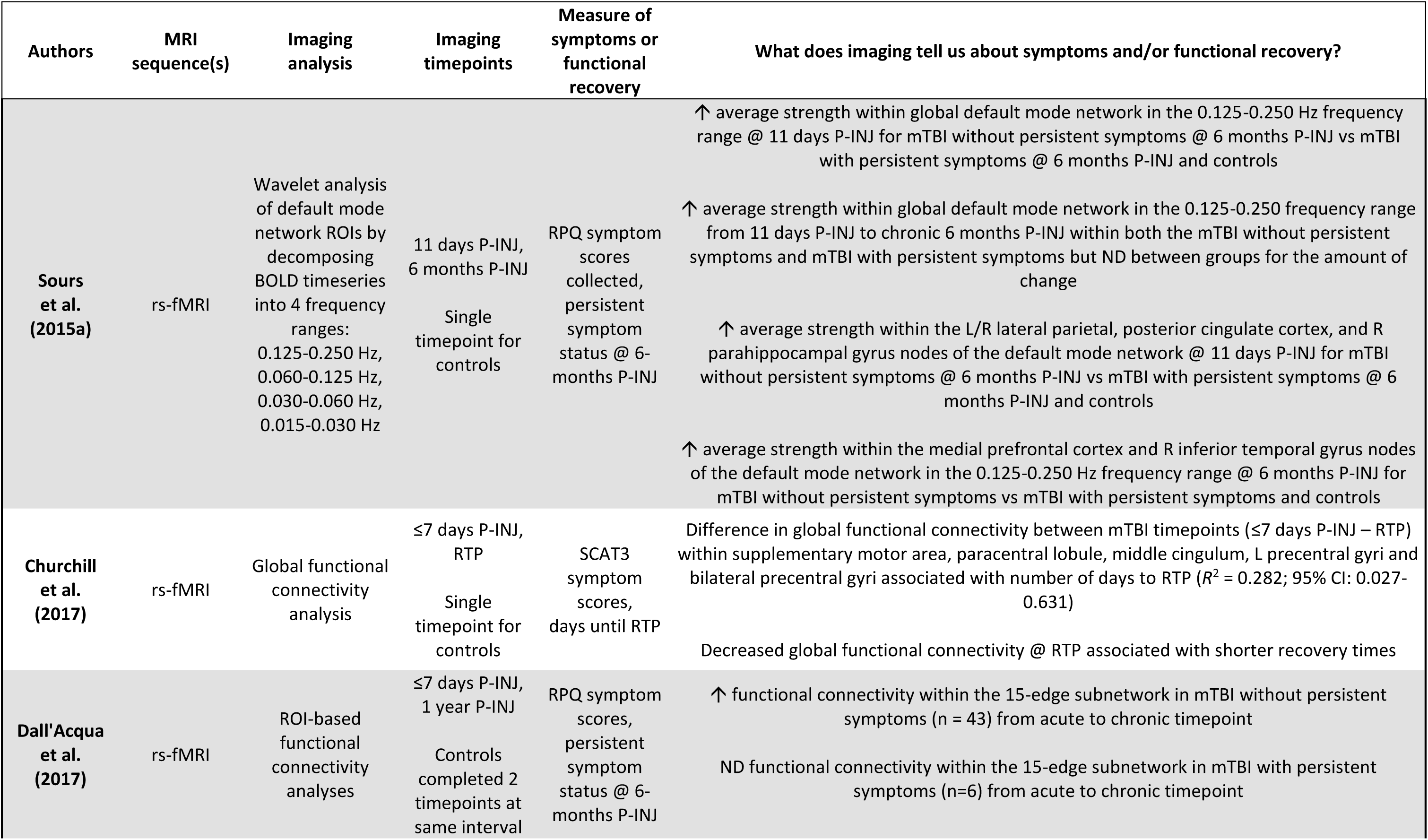
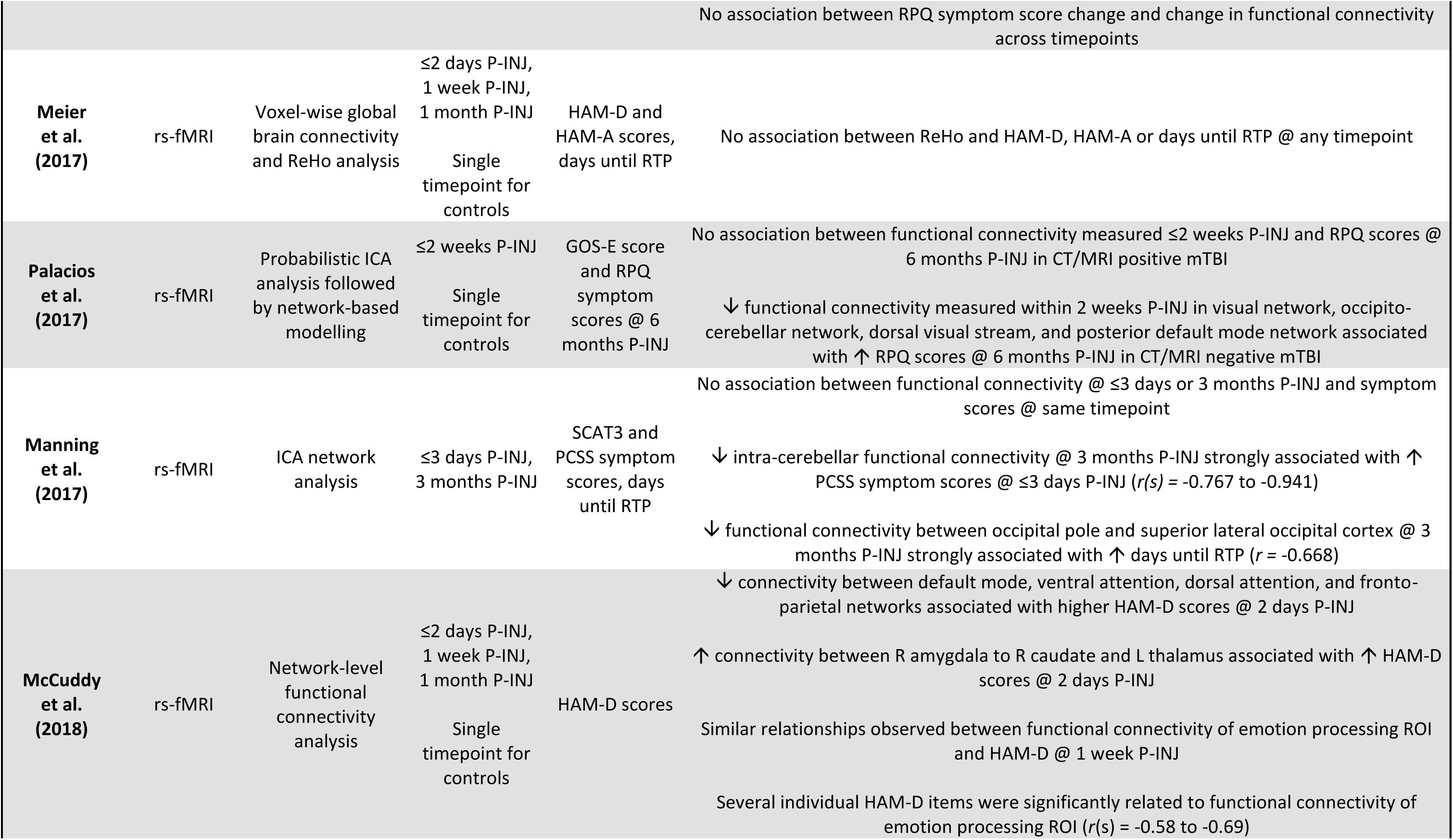
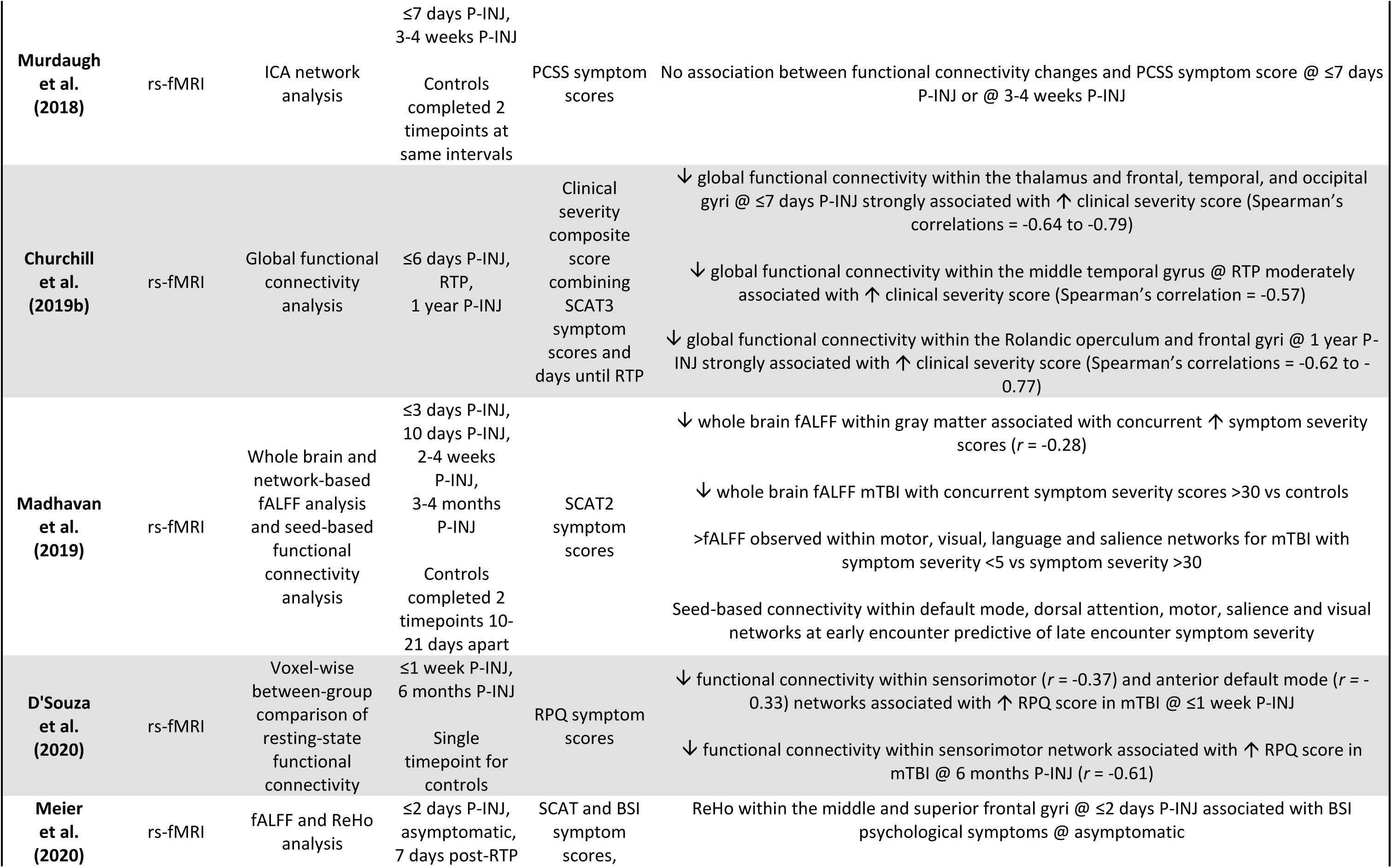
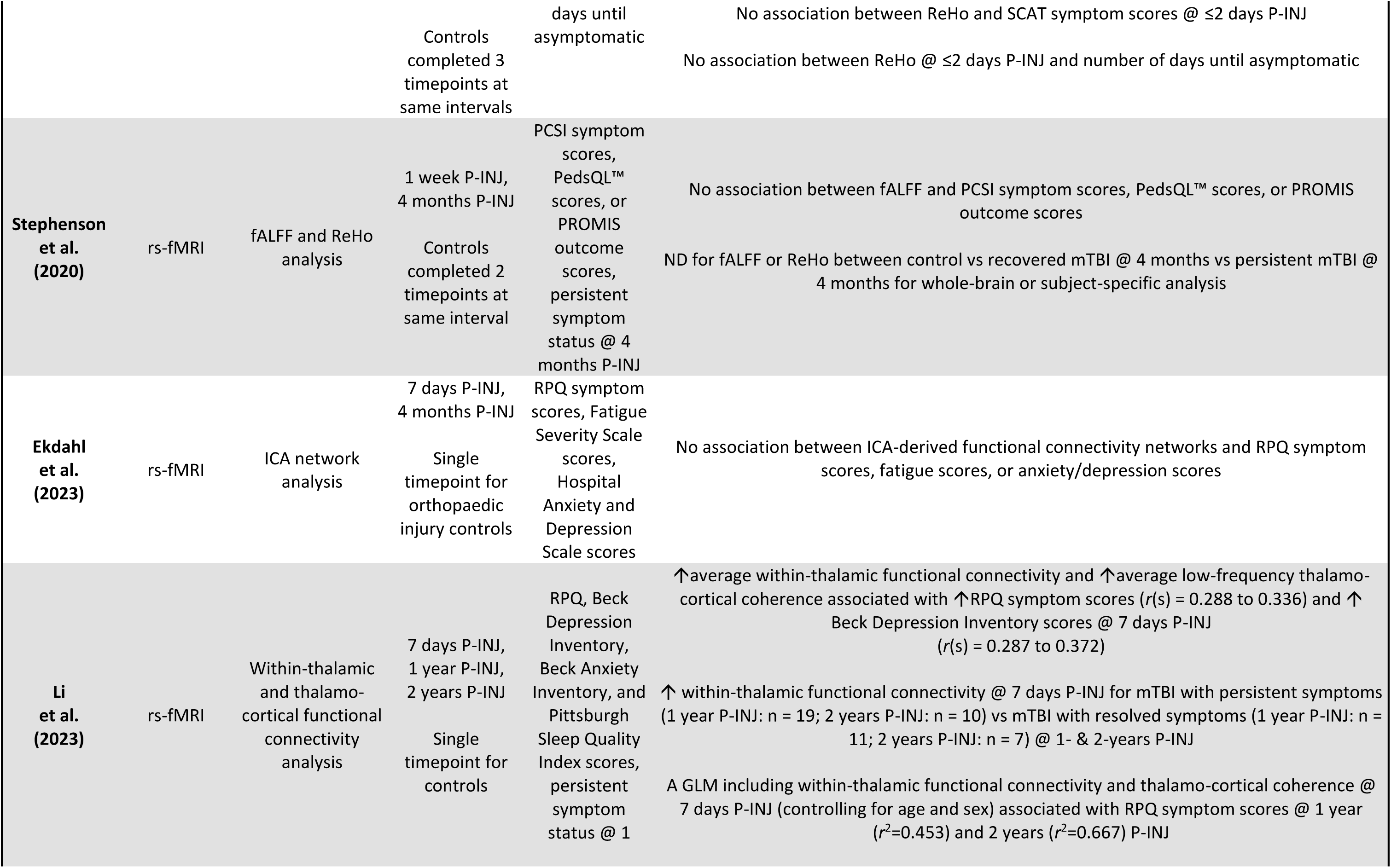
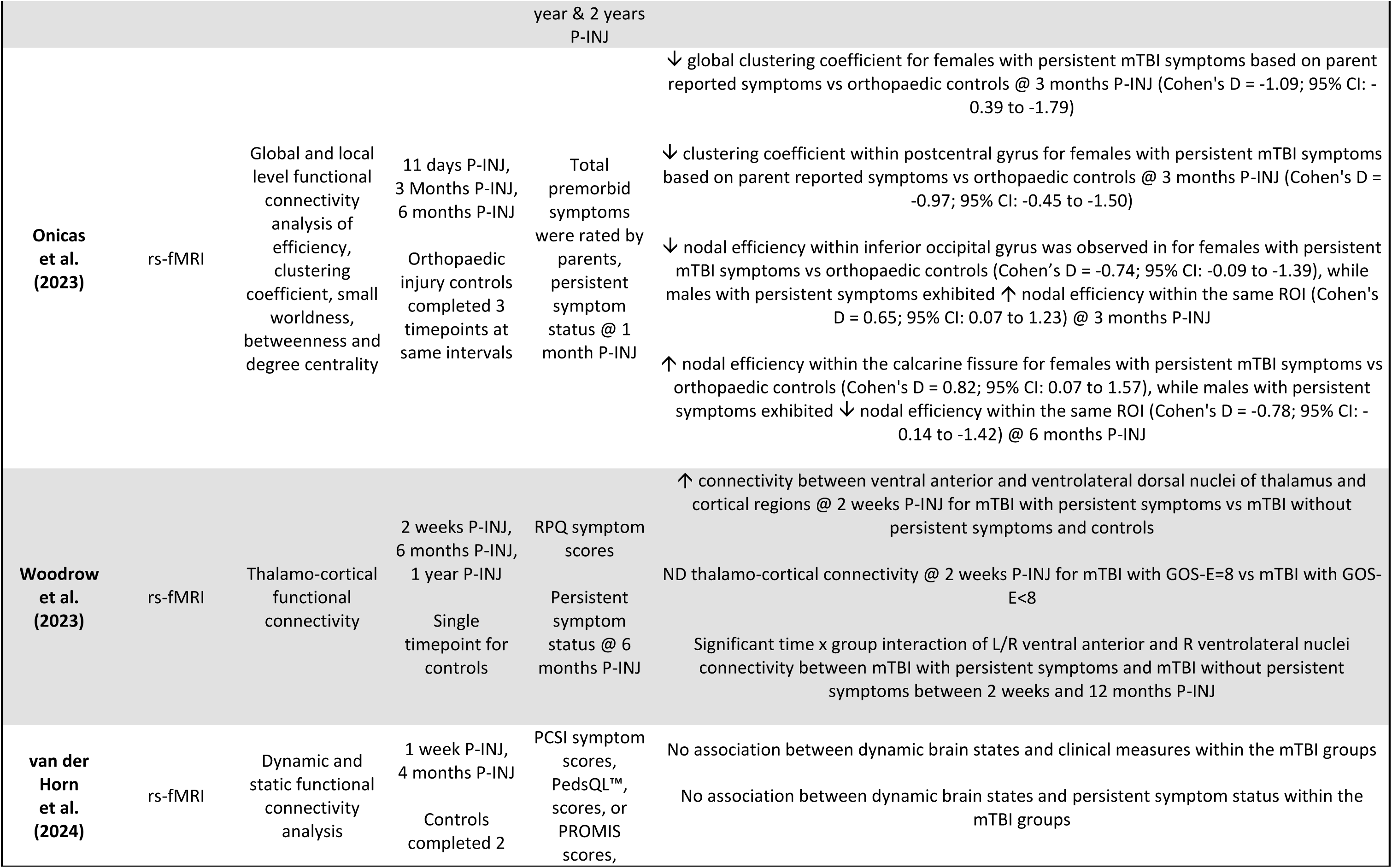
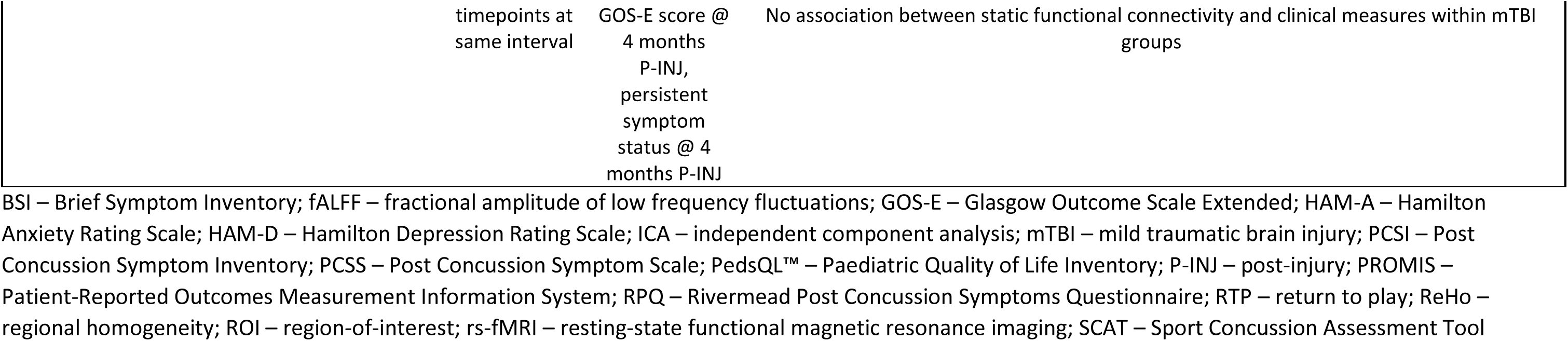
Results from studies using Blood-Oxygenation Level Dependent imaging.

Seven rs-fMRI studies explored network-specific functional connectivity using seed-to-voxel or independent component analysis (ICA) methods (Manning et al. 2017, Palacios et al. 2017, McCuddy et al. 2018, Murdaugh et al. 2018, Madhavan et al. 2019, D’Souza et al. 2020, Ekdahl et al. 2023). Three small studies (total n = 28, 30, and 43) observed no association between network-based functional connectivity and mTBI symptom scores (measured by PCSS, SCAT3, RPQ, Fatigue Severity Scale, or HADS) within seven days, or at one, three, or four months P-INJ (Manning et al. 2017, Murdaugh et al. 2018, Ekdahl et al. 2023). However, one study found that decreased functional connectivity at three months of P-INJ was associated with longer RTP times and greater acute symptom burden (measured by PCSS) (Manning et al. 2017). A larger study (total n = 94) reported reduced connectivity between default mode, ventral attention, dorsal attention, and fronto-parietal networks and increased connectivity between amygdala-caudate and amygdala-thalamus associated with higher symptoms scores (measured by HAM-D) at two days and one week P-INJ (McCuddy et al. 2018). D’Souza et al. (2020; total n = 120) described reduced functional connectivity within sensorimotor and anterior default mode networks and demonstrated a weak-moderate positive relationship with increased symptom scores (measured by RPQ) at seven days P-INJ. At six months P-INJ, reduced connectivity within the sensorimotor network was strongly associated with increased symptom scores (D’Souza et al. 2020). Two studies reported associations between acute network-based functional connectivity and greater symptom scores 1-6 months P-INJ. Madhavan et al. (2019; total n = 114) observed that acute seed-based connectivity within default mode, dorsal attention, motor, salience and visual networks predicted symptom severity (measured by SCAT) at 1-4 months P-INJ. The findings of Palacios et al. (2017; total n = 122) support this finding with results indicating that reduced functional connectivity within the visual, occipito-cerebellar, dorsal visual stream, and posterior default mode network were negatively associated with increased symptom scores (measured by RPQ) at six months P-INJ in mTBI patients with negative CT/conventional MRI findings.

Four studies evaluated voxel-wise global functional connectivity (Churchill et al. 2017, Churchill et al. 2019b, Onicas et al. 2023, van der Horn et al. 2024). One study (total n = 54) described differences in global functional connectivity from seven days P-INJ to RTP demonstrated a moderate-strong relationship with number of days to RTP. Specifically, decreases in global functional connectivity at RTP were associated with short recovery times (Churchill et al. 2017). Similarly, Churchill et al. (total n = 146) observed widespread acute reductions in global functional connectivity were strongly associated with worse acute symptom severity (measured by SCAT3) and longer RTP (Churchill et al. 2019b). The significant clusters were restricted to the middle temporal gyrus at RTP (Churchill et al. 2019b). A large study (total n = 376) reported no associations between dynamic brain states or static functional connectivity and symptom scores (measured by PCSI, Paediatric Quality of Life Inventory, or PROMIS) or recovery/persistent symptom status (van der Horn et al. 2024). Another study (total n = 264) observed large reductions in global clustering coefficient at three months P-INJ for females with persistent mTBI symptoms compared to orthopaedic injury controls (Onicas et al. 2023).

Voxel-wise regional homogeneity (ReHo) and/or fALFF (fractional amplitude of low-frequency fluctuations) were quantified in four studies (Meier et al. 2017, Madhavan et al. 2019, Meier et al. 2020, Stephenson et al. 2020). One study (total n = 114) found decreased fALFF within gray matter was weakly associated with increased symptom scores (measured by SCAT) and that mTBI patients with lower symptom scores demonstrated greater fALFF within the motor, visual, language, and salience networks compared to mTBI patients with high symptom scores (Madhavan et al. 2019). The remaining three studies (total n = 94, 174, and 293) reported no associations between ReHO or fALFF and symptom scores (measured by PCSI, Paediatric Quality of Life Inventory, PROMIS, HAM-A, HAM-D, or SCAT). Additionally, no differences in fALFF or ReHo were reported based on days until asymptomatic/RTP or symptomatic status (Meier et al. 2017, Meier et al. 2020, Stephenson et al. 2020).

Region-of-interest-based functional connectivity analysis was used in two studies (Dall’Acqua et al. 2017, Onicas et al. 2023). The first study (total n = 98) reported increased functional connectivity within a subnetwork from seven days to one year P-INJ for mTBI patients with resolved symptoms. However, there was no difference in connectivity within this subnetwork for patients with persistent symptoms. No associations were observed between the change in functional connectivity and symptom scores (measured by RPQ) between timepoints (Dall’Acqua et al. 2017). The other study (total n = 264) described large sex-based differences for nodal efficiency within the inferior occipital gyrus and calcarine fissure at three- and six-months P-INJ for paediatric mTBI patients with persistent symptoms compared to controls (Onicas et al. 2023).

Two studies investigated thalamo-cortical functional connectivity (Li et al. 2023, Woodrow et al. 2023). The first study (total n = 140) observed a weak-moderate positive correlation between increased intra-thalamic and thalamo-cortical connectivity and increased symptom scores (measured by RPQ and BSI) at seven days P-INJ (Li et al. 2023). Greater intra-thalamic connectivity was reported at seven days P-INJ for mTBI patients with persistent symptoms at 1-2 years P-INJ compared to patients with resolved symptoms. Intra-thalamic and thalamo-cortical connectivity at seven days P-INJ was strongly associated with increased symptom scores (measured by RPQ) at one year P-INJ (Li et al. 2023). The second study (total n = 184) also described increased thalamo-cortical connectivity at two weeks P-INJ for mTBI patients with persistent symptoms at six months P-INJ compared to patients with resolved symptoms and controls (Woodrow et al. 2023). However, there were no differences in thalamo-cortical connectivity at two weeks P-INJ for those with/without incomplete recovery (measured by GOS-E) at six months P-INJ (Woodrow et al. 2023).

A single study (total n = 63) performed wavelet analysis (Sours et al. 2015a). The authors described increased average strength in the 0.125-0.250 Hz range within default mode network, lateral parietal cortex, and posterior cingulate cortex observed at 10 days P-INJ for mTBI patients with resolved symptoms compared to patients with persistent symptoms at six months P-INJ and controls. There was no difference in the degree of change within the default mode network from 10 days to six months P-INJ for patients with/without persistent symptoms (Sours et al. 2015a).

#### 3.3.4 Arterial spin labelling imaging and functional recovery outcomes

Six studies described relationships between ASL-derived CBF and measures of mTBI symptoms and/or functional recovery (Meier et al. 2015a, Sours et al. 2015b, Stephens et al. 2018, Churchill et al. 2019b, Wang et al. 2019, Sicard et al. 2024). Across the six studies (Table 6), three different methods of ASL were used including: standard ASL (Meier et al. 2015a, Churchill et al. 2019b, Wang et al. 2019), pseudo-continuous ASL (Stephens et al. 2018), and pulsed ASL (Sours et al. 2015b, Sicard et al. 2024). The first study (total n = 44) reported a large reduction in CBF within dorsal midinsular cortex for mTBI patients who required more than two weeks to RTP compared to those who returned to play in less than two weeks (Meier et al. 2015a). Additionally, reduced CBF in this ROI at one month P-INJ was associated with simultaneously increased symptom scores (measured by HAM-A and HAM-D) (Meier et al. 2015a). Another study (total n = 56) observed increased CBF within the default mode network compared to task positive network in mTBI patients with resolved symptoms – but not patients with persistent symptoms – at one week, one month, and six months P-INJ. The authors also reported no differences in CBF for mTBI patients with/without persistent symptoms compared to controls at any timepoint (Sours et al. 2015b). In a small study (total n = 30), no differences in rCBF were described for mTBI patients with high or low symptom scores (measured by PCSS) compared to controls within two weeks P-INJ. However, increased rCBF within dorsal anterior cingulate cortex was observed at six weeks P-INJ in mTBI patients with physical symptoms vs controls (Stephens et al. 2018). A larger study (total n = 146) reported a moderate-strong negative association between acute CBF within the middle frontal, middle temporal, and superior parietal gyrus and acute symptom severity and RTP times (Churchill et al. 2019b). No associations between CBF and clinical outcomes were observed at RTP. Wang et al. (2019; total n = 48) noted reduced rCBF within the inferior parietal lobule and supramarginal gyrus at two days P-INJ was moderately negatively correlated with increased number of days until asymptomatic. The final study (total n = 99) observed increased adjusted perfusion within precuneus and reduced adjusted perfusion within superior frontal gyrus was associated with increased symptom scores (measured by HBI) at four days and one month P-INJ (Sicard et al. 2024). Reduced ROI-based adjusted perfusion was observed in a small subgroup (n = 10) of symptomatic mTBI patients compared to controls at one month P-INJ, while increased ROI-based adjusted perfusion was reported in asymptomatic patients relative to controls (Sicard et al. 2024)

**Table 6.** Results from studies using Arterial Spin Labelling imaging.

| Authors | MRI sequence(s) | Imaging analysis | Imaging timepoints | Measure of symptoms or functional recovery | What does imaging tell us about symptoms and/or functional recovery? |
| --- | --- | --- | --- | --- | --- |
| **Meier et al. (2015)** | ASL | Voxel-wise linear mixed-effects model to assess changes in CBF as a function of recovery in mTBI group | ≤2 days P-INJ, 1 week P-INJ, 1 month P-INJ<br>Single timepoint for controls | HAM-D and HAM-A, recovery outcome based on days to RTP: >14 days = poor outcome | ↓ CBF mTBI with poor outcome vs mTBI with good outcome within R dorsal midinsular cortex @ 1 month P-INJ (Cohen's D = 1.92)<br>CBF within R dorsal midinsular cortex @ 1 month P-INJ associated with T1 HAM-D total symptom score ($r = -0.64$)<sup>†</sup> and T1 HAM-A total symptom score ($r = -0.56$)<sup>†</sup> |
| **Sours et al. (2015b)** | PASL | CBF ratio between CBF maps extracted from default mode and task positive networks | 1 week P-INJ, 1 month P-INJ, 6 months P-INJ<br>Single timepoint for controls | RPQ symptom scores, persistent symptom status | ↑ CBF in the default mode network vs task positive network for mTBI without persistent symptoms @ all timepoints<br>ND CBF in the default mode network vs task positive network for mTBI with persistent symptoms @ all timepoints<br>ND CBF in default mode network, task positive network or CBF ratio for mTBI with persistent symptoms and/or mTBI without persistent symptoms vs controls @ any timepoint |
| **Stephens et al. (2018)** | PCASL | Whole-brain and ROI-based analysis of rCBF maps | ≤2 weeks P-INJ, 6 weeks P-INJ<br>Single timepoint for controls | PCSS symptom scores, high vs low symptom subgroups @ <2 weeks P-INJ, persistent symptom status @ 6 weeks P-INJ | ND in rCBF for high symptom vs low symptom mTBI (physical, emotional, cognitive, sleep-related categories) vs controls @ ≤2 weeks P-INJ<br>ND rCBF within L dorsal anterior cingulate cortex for mTBI without presence of physical symptoms vs controls @ ~6 weeks P-INJ<br>↑ rCBF within L dorsal anterior cingulate cortex for mTBI with presence of physical symptoms vs controls @ ~6 weeks P-INJ |
| **Churchill et al. (2019b)** | ASL | Voxel-wise analysis of CBF maps | ≤6 days P-INJ, RTP, 1 year P-INJ | Clinical severity composite score combining SCAT3 | ↓ CBF within middle frontal, middle temporal, and superior parietal gyrus @ ≤7 days P-INJ strongly associated with ↑ clinical severity score (Spearman's correlations = -0.63 to -0.78)<br>No association between CBF and clinical severity score @ RTP |
|  |  |  |  | symptom scores and days until RTP | ↑ CBF within inferior frontal gyrus @ 1 year P-INJ strongly associated with ↑ clinical severity score (Spearman's correlations = 0.73 to 0.75) |
| **Wang et al. (2019)** | ASL | Voxel-wise analysis of CBF maps | ≤2 days P-INJ<br>Single timepoint for controls | SCAT3 symptom scores, days until asymptomatic | ↓ rCBF within R inferior parietal lobule ($r = -0.47$) and R supramarginal gyrus ($r = -0.44$) @ ≤2 days P-INJ associated with ↑ number of days until asymptomatic |
| **Sicard et al. (2024)** | PASL | Z-score perfusion maps were used to identify voxels of abnormal hyperperfusion or hypoperfusion | ≤4 days P-INJ, 1 month P-INJ<br>Single timepoint for orthopaedic injury controls | HBI symptom scores, symptomatic status | ↑ adjusted perfusion within L precuneus and ↓ adjusted perfusion within L superior frontal gyrus associated with ↑ HBI symptom scores in mTBI @ both timepoints<br>↓ adjusted perfusion within R cerebellum and R lingual gyrus symptomatic mTBI (n = 10) vs orthopaedic control @ 1 month P-INJ<br>↑ adjusted perfusion within L medial orbitofrontal cortex, R middle frontal gyrus and bilateral caudate and ↓ adjusted perfusion within bilateral calcarine asymptomatic mTBI (n = 59) vs orthopaedic control @ 1 month P-INJ |

<sup>†</sup> - no multiple comparisons correction reported; ASL – Arterial Spin Labelling; CBF – cerebral blood flow; rCBF – relative cerebral blood flow; HAM-A – Hamilton Anxiety Rating Scale; HAM-D – Hamilton Depression Rating Scale; HBI – Health and Behaviour Inventory; mTBI – mild traumatic brain injury; PASL – Pulsed Arterial Spin Labelling; PCASL – Pseudo-Continuous Arterial Spin Labelling; PCSS – Post Concussion Symptom Scale; P-INJ – post-injury; RPQ – Rivermead Post Concussion Symptoms Questionnaire; ROI – region-of-interest; SCAT – Sport Concussion Assessment Tool

## 4. Discussion

This review collated evidence on the utility of MRI techniques used during the acute phase following mTBI to detect pathophysiological correlates of symptom burden and functional recovery. Due to heterogeneity in study designs and reporting, meta-analysis was not possible. Instead, a qualitative synthesis of key findings is provided, emphasizing results replicated in multiple studies. In cases of conflicting results, greater weight is given to findings from large multi-site studies.

### 4.1 Modality-specific key findings

Pathoanatomic features of mTBI, evaluated using structural and SWI methods, show limited association with clinical outcomes. Similar incidence and prevalence of pathoanatomic features were observed in mTBI participants and controls in multiple studies (Studerus-Germann et al. 2018, Mayer et al. 2020a). In a large multi-site study of paediatric mTBI, Mayer et al. found no association between pathoanatomic MRI findings and symptom reports or quality of life (Mayer et al. 2020a). Similarly, cortical thickness and volumes derived from T1-weighted imaging show limited association with persistent symptom status (Bobholz et al. 2021, Mayer et al. 2023a). However, changes in volume within limbic and subcortical structures from acute to six months post-injury were greater in mTBI patients with persistent symptoms and moderately correlated with worse symptoms and GOS-E scores (Zhuo et al. 2021, Xu et al. 2023).

In microstructural investigations of mTBI, there appears to be limited evidence suggesting a direct association between TBSS and voxel-wise DTI metrics and symptom burden within the same timepoint. Conversely, acute changes in white matter microstructure following mTBI, assessed through TBSS, voxel-wise, and tract-based methods, were found to be associated with or predictive of symptom burden and time to recovery at follow-up timepoints (e.g. 3-6 months P-INJ; (Yuh et al. 2014, Churchill et al. 2019b, Ware et al. 2020, Wu et al. 2020, Mayer et al. 2022, Palacios et al. 2022, Li et al. 2023, Pinto et al. 2023)). Specific microstructural metrics associated with worse symptom burden and recovery varied across studies, with some reporting increased MD (Ware et al. 2020, Wu et al. 2020, Pinto et al. 2023), AD (Palacios et al. 2022), RD (Li et al. 2023), reduced FA (Yuh et al. 2014, Ware et al. 2020, Mayer et al. 2022, Pinto et al. 2023), and increased ODI, V*_IC_*, and V*_ISO_* (Mayer et al. 2022). Interestingly, two TBSS studies also found that greater acute symptom burden was associated with reduced FA and increased MD and RD at follow-up 3-6 months P-INJ (Yin et al. 2019, Wu et al. 2020).

The evidence regarding concurrent correlations between functional connectivity and symptom burden is inconsistent within the literature. Overall, rs-fMRI-derived functional connectivity may capture weak-moderate associations with symptom burden at the same timepoint (McCuddy et al. 2018, Madhavan et al. 2019, D’Souza et al. 2020, Li et al. 2023). These associations were reported using network-based and voxel-wise fALFF analyses. Similar to microstructural findings, acute alterations in functional connectivity were associated with worse symptom burden and recovery outcomes at follow-up timepoints (Palacios et al. 2017, Churchill et al. 2019b, Madhavan et al. 2019, Li et al. 2023, Woodrow et al. 2023). Specifically, reduced functional connectivity within the default mode, visual, and task-positive networks (Palacios et al. 2017, Madhavan et al. 2019) and increased intra-thalamic/thalamo-cortical connectivity (Li et al. 2023, Woodrow et al. 2023) were associated with worse clinical outcomes at follow-up.

Studies examining ASL-derived CBF following mTBI report varied findings. Four studies suggest that changes in CBF may be linked to a greater symptom burden and longer recovery times. However, the direction of CBF changes and specific ROI where changes were observed varied across these studies (Meier et al. 2015a, Churchill et al. 2019b, Wang et al. 2019, Sicard et al. 2024). Additionally, only 10% of the studies in this review investigated the relationship between CBF and mTBI clinical outcomes, indicating a need for further exploration in this area.

### 4.2 Neuroimaging insights into recovery

Appraisal of the neuroimaging literature has provided valuable insights into mTBI recovery outcomes and underlying neural mechanisms. Notably, three anatomical structures—the thalamus, cingulate cortex, and superior longitudinal fasciculus (SLF) —were each associated with poor recovery outcomes in five or more studies (Figure 3). Reduced thalamic volume, intra-thalamic functional connectivity, thalamo-cortical functional connectivity, decreased global functional connectivity within the thalamus, diffusion changes within anterior thalamic radiation, and changes in clustering coefficient within the thalamus corresponded with persistent symptom status (McCuddy et al. 2018, Churchill et al. 2019b, Zhuo et al. 2021, Ware et al. 2022, Li et al. 2023, Ware et al. 2023, Woodrow et al. 2023). Greater susceptibility and diffusion changes within the SLF were related to persistent symptom status, symptom severity, and increased RTP time (Meier et al. 2016, Manning et al. 2017, Evans et al. 2018, Churchill et al. 2019a, Palacios et al. 2022, Ware et al. 2022, Brown et al. 2023).

**Figure 3.**
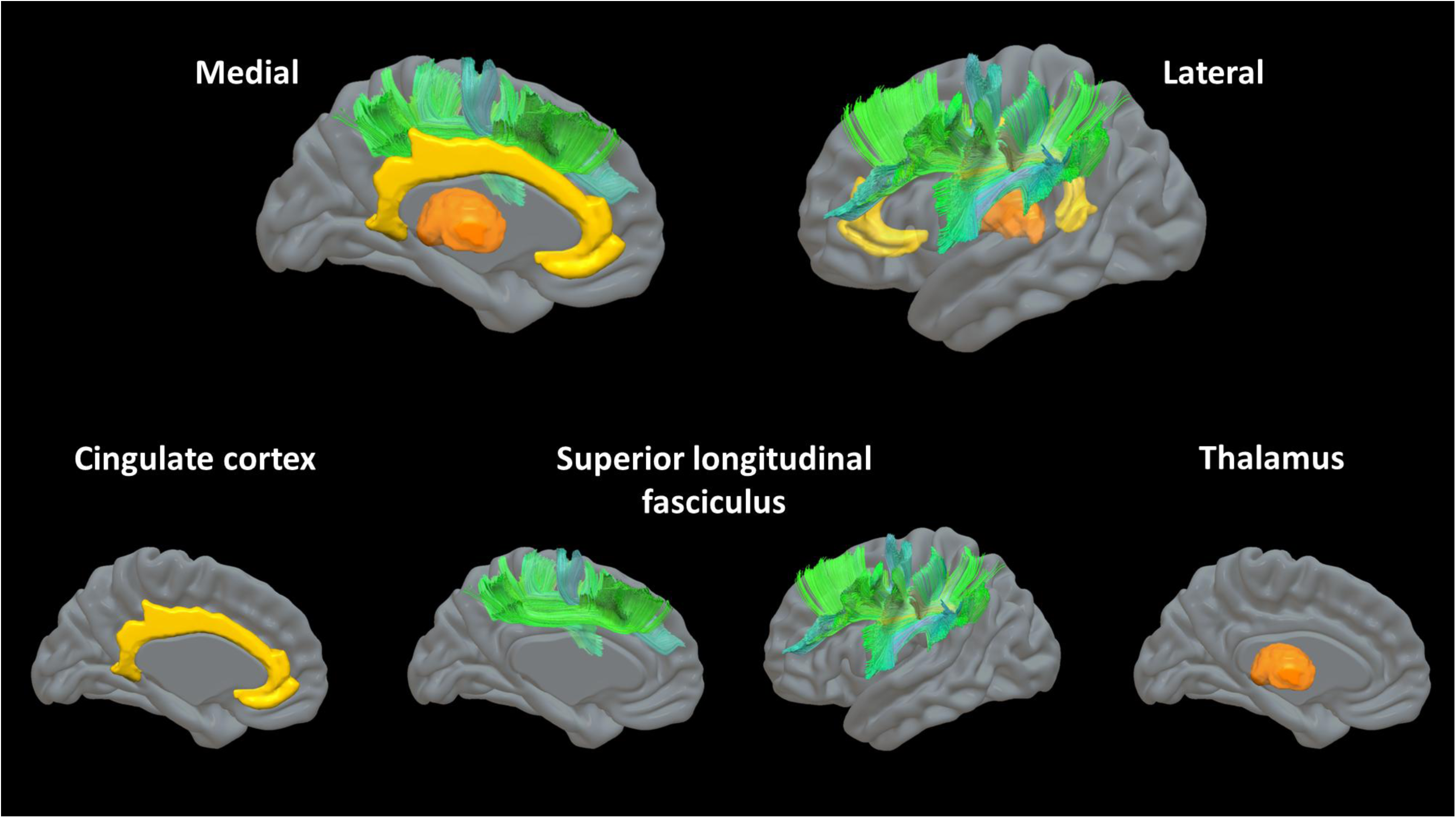
Structures-of-interest associated with mTBI recovery outcomes with the cingulate cortex shown in yellow, the thalamus shown in orange, and the superior longitudinal fasciculus (SLF) shown in blue-green. Visualization of the cingulate cortex and thalamus are based on the Harvard-Oxford Cortical and Subcortical Structural Atlas (Desikan et al. 2006); while visualization of the SLF is based on the HCP1065 Population-Averaged Tractography Atlas (Yeh 2022).

Changes in functional connectivity, fALFF, and CBF within the anterior, middle, and/or posterior cingulate cortex were also associated with symptom burden and persistent symptom status (Sours et al. 2015a, Sours et al. 2015b, Churchill et al. 2017, Palacios et al. 2017, McCuddy et al. 2018, Stephens et al. 2018, Madhavan et al. 2019, Xu et al. 2023). These observations highlight two impactful trends. First, these structures serve as crucial hubs for integrating and relaying cognitive, sensory, and emotional stimuli throughout the brain. The current literature suggests that greater mTBI-related disruption to these central hubs indicates a higher likelihood of more severe symptoms and longer recovery outcomes. Second, disruptions related to mTBI in each of these structures were observed using multiple imaging modalities, indicating the multi-scale impact of injury on these structures and its implications for recovery. These findings suggest that multiscale disruptions to central hubs such as the thalamus, SLF, or cingulate cortex may underpin the heterogeneity of mTBI symptoms and recovery outcomes. Rather than attributing variation in outcomes to damage in localized regions with more specific functions, injury to these multifunctional hubs – critical for integrating diverse cognitive, sensory, and emotional processes – plays a significant role in mTBI outcomes.

Several studies used follow-up clinical data to stratify mTBI patients based on persistent symptom status (evaluated at least six months P-INJ). They assessed their acute imaging findings to identify initial differences in recovery trajectories. These studies suggest divergent trends in acute neuroimaging markers between patients who develop persistent symptoms and those who recover fully (Sours et al. 2015a, Palacios et al. 2022, Ware et al. 2022, Li et al. 2023, Ware et al. 2023, Woodrow et al. 2023). For instance, acute increases in FA were reported in paediatric mTBI patients who did not develop persistent symptoms, whereas reduced FA was associated with persistent symptoms (Ware et al. 2022, Li et al. 2023). Additionally, acute differences in structural and functional connectivity were frequently observed in patients with persistent symptoms compared to controls. At the same time, those asymptomatic at the follow-up showed no acute differences (Sours et al. 2015a, Ware et al. 2022, Li et al. 2023, Ware et al. 2023, Woodrow et al. 2023). Whether these differences are attributable to individual and/or injury-related factors remains unclear. Ware et al. and Onicas et al. used data from the A-CAP initiative to explore whether differential injury responses, measured by neuroimaging in paediatric mTBI patients who developed persistent symptoms, were influenced by sex and age. The authors reported reduced ROI-based measures of structural and functional connectivity in females who developed persistent symptoms compared to controls (i.e. reduced clustering coefficient within the putamen and reduced nodal efficiency within the inferior occipital gyrus). In contrast, males with persistent symptoms showed the opposite effect within the same ROI (Onicas et al. 2023, Ware et al. 2023). Furthermore, Ware et al. also observed reduced betweenness centrality associated with the supramarginal gyrus and putamen in younger children with persistent symptoms compared to controls (Ware et al. 2023). Whereas the opposite effect was seen in older children who developed persistent symptoms (Ware et al. 2023).

### 4.3 Considerations for future studies

This review has identified five recommendations that should be considered in future studies to strengthen the mTBI neuroimaging literature. First, it is *strongly* recommended that all future mTBI neuroimaging studies collect and report a measure of symptom burden and/or functional recovery. The primary reason for excluding 124 studies during the screening phase of this review was the absence of a functional recovery outcome measure (Figure 1). An additional 17 studies were excluded because, although they included a recovery measure, they did not report any findings relating the recovery measure to neuroimaging results. Considering the expense and effort of neuroimaging research, failing to simultaneously acquire data that elucidate the clinical significance of neuroimaging findings represents a missed opportunity. There are few barriers to the inclusion of these measures (i.e. GOS-E or PCSS) as they are easy, fast (≤10 minutes), and free to administer. The current understanding of the heterogeneity of mTBI symptomology and recovery, as well as the neuroimaging methods best equipped to explain these complexities, could be significantly advanced if these measures were consistently included and analysed.

Second, investigators should use clinical data to identify meaningful subgroups/phenotypes of mTBI patients to guide neuroimaging analysis. This review highlights the value of this approach through the insights gained by analysing acute imaging data stratified by persistent symptom status at follow-up. However, this methodology can be extended to explore neurobiological correlates of different symptom-driven phenotypes within mTBI patients. For example, Cai et al. reported reduced global and tract-based AD in mTBI patients with neuropsychiatric distress compared to those with emotional resilience at both acute and follow-up timepoints (Cai et al. 2023). A subset of studies used RPQ, HBI, or PCSS symptom subscales to specifically analyse cognitive, emotional, somatic, and/or sleep disturbance phenotypes and their relation to neuroimaging findings (Evans et al. 2018, Stephens et al. 2018, Studerus-Germann et al. 2018, Baker et al. 2020, Ware et al. 2020, Zhuo et al. 2021, Ware et al. 2022, Woodrow et al. 2023). Aggregating these findings is challenging at this stage due to differences in the modalities of neuroimaging acquired and analyses performed. Nonetheless, this method should be explored in further detail to understand whether specific regions-, networks-, or tracts-of-interest underpin the various symptom reporting patterns and patient phenotypes observed in clinical practice.

Third, authors should accompany their tests of statistical significance with standardised measures of effect. Less than half of studies in this review reported the magnitude of the observed difference/association reported in their results. Seventeen studies provided a correlation coefficient or coefficient of determination (Yuh et al. 2014, Meier et al. 2015a, Meier et al. 2016, Churchill et al. 2017, Manning et al. 2017, Koch et al. 2018, McCuddy et al. 2018, Churchill et al. 2019b, Madhavan et al. 2019, Wang et al. 2019, Yin et al. 2019, D’Souza et al. 2020, Wu et al. 2020, Zhuo et al. 2021, Gugger et al. 2023, Li et al. 2023, Xu et al. 2023), five studies stated Cohen’s D effect estimates (Meier et al. 2015a, Ware et al. 2022, Cai et al. 2023, Onicas et al. 2023, Ware et al. 2023), and only two studies reported odds ratios (Yuh et al. 2014, Palacios et al. 2022). Understanding the magnitude of difference/association of statistically significant findings is essential to advancing our understanding of mTBI symptomology and recovery.

Fourth, future efforts are encouraged to extend beyond simply testing for group differences by exploring the predictive capability of their observed findings. For instance, can we use neuroimaging data to accurately *predict* who is likely to have persistent symptoms versus those who are not, or can we *classify* individuals with a cognitive symptom phenotype versus emotional or somatic? By incorporating additional analyses to explore these questions, we can advance our understanding of major clinical issues in mTBI, such as identifying individuals needing targeted treatment and determining the specific treatments required to address their underlying issues. We recommend interested readers refer to Fleck et al. (2021), Gugger et al. (2023), Mayer et al. (2022), Palacios et al. (2022), and Pinto et al. (2023) as good examples of incorporating these analytical frameworks.

Finally, research groups may consider adopting integrative analytical methods to understand how multiple neuroimaging modalities jointly explain or predict differences/associations in mTBI symptom burden and recovery. In this review, several studies reported results for multiple imaging modalities in the same article, but analysed each modality in isolation, which is consistent with a reductionist approach (Churchill et al. 2017, Dall’Acqua et al. 2017, Murdaugh et al. 2018, Churchill et al. 2019b, Li et al. 2023). Here, we challenge the community to embrace a biological systems approach to understanding the heterogeneity of mTBI (Ahn et al. 2006). Rather than viewing each imaging modality as an independent aspect of the brain’s response to injury, consider each modality a partial representation of multi-component covariation across the entire brain (Stone et al. 2020, Avants et al. 2021). This review has demonstrated how qualitatively integrating findings across multiple modalities helped identify the thalamus, SLF, and cingulate cortex as structures-of-interest related to mTBI recovery outcomes. Given the volume and complexity of these datasets, quantitative integrative analysis may be well suited for thoughtful applications of machine learning, such as featureless neural networks. This approach would be particularly fitting for research questions interested in exploring the accuracy of multi-modality neuroimaging datasets to predict mTBI symptomology and recovery trajectories at the expense of interpretability of features contributing to predictive performance. In contrast, integrative statistical designs enable dimensionality reduction of joint relationships within the data into interpretable components that can be used in standard null-hypothesis testing or machine learning algorithms (Stone et al. 2020, Avants et al. 2021). Application of this approach has yielded novel insights into the multimodal effects of low-level blast-exposure on brain health in military members (Stone et al. 2020, Stone et al. 2023). Overall, migration towards methodological and analytical frameworks that embrace the complexity of mTBI may unlock our understanding of the variability in patient experience and outcomes.

## 5. Limitations

A limitation of this review is that included studies were not constrained by specific MRI acquisition parameters or post-processing software. During the screening process, it became clear that there were considerable inconsistencies, particularly in DTI studies, which used different spatial resolutions, b-values, single-versus multi-shell data, and varied approaches to tractography. Previous research has demonstrated that differences in such aspects can impact study findings. Whilst this has been the motivation behind efforts to harmonize acquisitions and processing across multiple MRI sites in large scale studies, this is not a trivial task and requires advanced deep learning techniques which are beyond the scope of the current review (Liu and Yap 2024, Marzi et al. 2024). It is worth noting that many of the multi-site studies included in this review deployed *ComBat* harmonization to account for these challenges (Fortin et al. 2018). Another limitation is that the review did not explicitly separate findings by participant age. This is important to acknowledge as 18 studies included samples with a mean age <18 years and Ware et al. highlighted differences in the relationship between imaging findings and recovery outcomes based on maturation status (Ware et al. 2022). Both limitations are important to consider when interpreting the state of the evidence.

Nevertheless, a key strength of this review is the consistency of trends observed across studies, despite differences in acquisition methods, processing techniques, and participant characteristics. For example, the relationship between mTBI-related changes in thalamic structure and function and recovery outcomes was identified in children, young adults, and adults, whether the sample consisted of athletic populations or patients from emergency departments. These trends emerged despite substantial variability in imaging acquisition and analysis methods. This consistency suggests that certain common effects of mTBI may influence recovery outcomes across a range of factors, including injury mechanism, age, and other methodological differences.

## 6. Conclusions

In conclusion, longitudinal studies reveal that acute neuroimaging markers differ between mTBI patients who develop persistent symptoms versus those who recover, with age and sex potentially influencing neuroimaging findings. Although structural MRI findings show limited correlation with mTBI clinical outcomes, acute changes in white matter and functional connectivity are more strongly associated with symptom burden and functional recovery outcomes. Across multiple imaging modalities, disruptions to multi-functional hubs such as the thalamus, cingulate cortex, and SLF were associated with increased symptom burden and worse recovery outcomes. Finally, this review highlights the value of integrating measures of symptom burden and functional recovery within neuroimaging investigations to understand mTBI and the need to make this standard practice in future research.

## Ethics approval

Not applicable

## CRediT authorship contribution statement

**Joshua P. McGeown:** Conceptualization, Methodology, Investigation, Data curation, Formal analysis, Visualization, Writing – original draft. **Mangor Pedersen:** Conceptualization, Methodology, Formal analysis, Writing – review and editing. **Remika Mito:** Investigation, Data curation, Formal analysis, Writing – review and editing. **Alice Theadom:** Writing – review and editing. **Jerome Maller:** Writing – review and editing. **Paul Condron:** Writing – review and editing. **Samantha Holdsworth:** Project administration, Conceptualization, Writing – review and editing.

## Disclosure of generative AI and AI-assisted technologies in the writing process

During the preparation of this work the author(s) used ChatGPT v3.5 in order to improve language and readability. After using this tool/service, the author(s) reviewed and edited the content as needed and take(s) full responsibility for the content of this publication.

## Abbreviations

AD: axial diffusivity
ACRM: American Congress of Rehabilitation Medicine
ASL: arterial spin labelling
AUC: area under the curve
BSI: Brief Symptom Inventory
CBF: cerebral blood flow
CDC: Centers for Disease Control and Prevention
CISG: Concussion in Sport Group
DKI: diffusion kurtosis imaging
DTI: diffusion tensor imaging
FA: fractional anisotropy
fALFF: fractional amplitude of low frequency fluctuations
FLAIR: fluid attenuated inversion recovery
GCS: Glasgow Coma Score
GOS-E: Glasgow Outcome Scale Extended
HAM-A: Hamilton Anxiety Rating Scale
HAM-D: Hamilton Depression Rating Scale
HBI: Health and Behaviour Inventory
ICA: independent component analysis
IQR: interquartile range
KAX: axial kurtosis
KFA: kurtosis fractional anisotropy
KRAD: radial kurtosis
LOC: loss of consciousness
MD: mean diffusivity
MK: mean kurtosis
mTBI: mild traumatic brain injury
NCAA-DOD CARE: National Collegiate Athlete Association and U.S. Department of Defense Concussion Assessment, Research, and Education Consortium
ND: no difference
NODDI: neurite orientation dispersion and density imaging
NS: not specified
ODI: orientation dispersion index
PASL: pulsed arterial spin labelling
PCASL: pseudo-continuous arterial spin labelling
PCSI: Post Concussion Symptom Inventory
PCSS: Post Concussion Symptom Scale
PedsQL™: Paediatric Quality of Life Inventory
P-INJ: post-injury
PROMIS: Patient-Reported Outcomes Measurement Information System
QSM: quantitative susceptibility mapping
rCBF: relative cerebral blood flow
RD: radial diffusivity
ReHo: regional homogeneity
ROI: region-of-interest
RPQ: Rivermead Post Concussion Symptoms Questionnaire
rs-fMRI: resting-state functional magnetic resonance imaging
RTP: return to play
SCAT: Sport Concussion Assessment Tool
SWI: susceptibility weighted imaging
TBSS: tract based spatial statistics
T1w: T1 weighted
T2w: T2 weighted
V*_IC_*: intracellular volume
V*_ISO_*: isotropic volume
WHO: World Health Organization

